# Quantifying the impact of immune history and variant on SARS-CoV-2 viral kinetics and infection rebound: a retrospective cohort study

**DOI:** 10.1101/2022.01.13.22269257

**Authors:** James A. Hay, Stephen M. Kissler, Joseph R. Fauver, Christina Mack, Caroline G. Tai, Radhika M. Samant, Sarah Connolly, Deverick J. Anderson, Gaurav Khullar, Matthew MacKay, Miral Patel, Shannan Kelly, April Manhertz, Isaac Eiter, Daisy Salgado, Tim Baker, Ben Howard, Joel T. Dudley, Christopher E. Mason, Manoj Nair, Yaoxing Huang, John DiFiori, David D. Ho, Nathan D. Grubaugh, Yonatan H. Grad

**Affiliations:** Harvard T.H. Chan School of Public Health, Boston, MA, USA; Yale School of Public Health, New Haven, CT, USA; University of Nebraska Medical Center, Omaha, NE, USA; IQVIA, Real World Solutions, Durham, NC, USA; Duke Center for Antimicrobial Stewardship and Infection Prevention, Durham, NC, USA; TEMPUS Labs, Chicago, IL, USA; Columbia University Vagelos College of Physicians and Surgeons, New York, NY, USA; Hospital for Special Surgery, New York, NY, USA; National Basketball Association, New York, NY, USA

## Abstract

**Background:** The combined impact of immunity and SARS-CoV-2 variants on viral kinetics during infections has been unclear.

**Methods:** We characterized 2,875 infections from the National Basketball Association occupational health cohort identified between June 2020 and January 2022 using serial RT-qPCR testing. Logistic regression and semi-mechanistic viral RNA kinetics models were used to quantify the effect of variant, symptom status, age, infection history, vaccination and antibody titer to founder SARS-CoV-2 strain on the duration of potential infectiousness and overall viral kinetics. The frequency of viral rebounds was quantified under multiple cycle threshold (Ct) value-based definitions.

**Results:** Among individuals detected partway through their infection, 51.0% (95% credible interval [CrI]: 48.2-53.6%) remained potentially infectious (Ct<30) five days post detection, with small differences across variants and vaccination history. Only seven viral rebounds (0.7%; N=999) were observed, with rebound defined as 3+ days with Ct<30 following an initial clearance of 3+ days with Ct≥30. High antibody titers against the founder SARS-CoV-2 strain predicted lower peak viral loads and shorter durations of infection. Among Omicron BA.1 infections, boosted individuals had lower pre-booster antibody titers and longer clearance times than non-boosted individuals.

**Conclusions:** SARS-CoV-2 viral kinetics are partly determined by immunity and variant but dominated by individual-level variation. Since booster vaccination protects against infection, longer clearance times for BA.1-infected, boosted individuals may reflect a less effective immune response, more common in older individuals, that increases infection risk and reduces viral RNA clearance rate. The shifting landscape of viral kinetics underscores the need for continued monitoring to optimize isolation policies and to contextualize the health impacts of therapeutics and vaccines.

**Funding:** Supported in part by CDC contract 200-2016-91779, Emergent Ventures at the Mercatus Center, the Huffman Family Donor Advised Fund, the MorrisSinger Fund, the National Basketball Association, and the National Basketball Players Association.

## Introduction

The viral kinetics of SARS-CoV-2 underlie the epidemiology of COVID-19 and the policies surrounding infection control. The amount and duration of viral shedding influences infectiousness (Ke et al., 2021, 2022; Marc et al., 2021; Marks et al., 2021; Puhach et al., 2022; Sun et al., 2021) and the duration of test positivity affect isolation policies, test recommendations, and clinical care guidelines.(Hellewell et al., 2021; Kissler, Fauver, Mack, Olesen, et al., 2021; Larremore et al., 2021; Mack et al., 2022; Néant et al., 2021; Quilty et al., 2021; Singanayagam et al., 2022) Descriptions of viral kinetics are also important for establishing baselines to measure the effectiveness of antiviral drugs. For example, rebounds of viral RNA concentrations and symptoms have been observed after antiviral treatment, but it has been unclear to what extent such rebounds also occur in the absence of drug.(Boucau, Uddin, et al., 2022; Charness et al., 2022) Most longitudinal viral kinetics studies pre-date the emergence of the BA.1 lineage, which features dramatic antigenic divergence from prior lineages, as well as the rollout of third and fourth vaccine doses.(Lusvarghi et al., 2022; Straten et al., 2022) Early findings on viral kinetics therefore need to be updated to account for extensive and heterogeneous immune experience across the population.(Cevik et al., 2021; Kissler, Fauver, Mack, Tai, et al., 2021)

To characterize the viral kinetics of SARS-CoV-2 infection, including rebounds, for the Delta and Omicron (BA.1 lineages BA.1.1529 and BA.1.1) variants in symptomatic and asymptomatic individuals with varied vaccination and infection histories, we measured viral RNA levels using densely-sampled RT-qPCR tests from 2,875 SARS-CoV-2 infections, each taken by combined anterior nares and oral swabs, that occurred between 7^th^ July, 2020, and 26^th^ January, 2022, prior to the detection of BA.2.12.1, BA.4 and BA.5, or the regular detection of BA.2, in this cohort. As a proxy for immune response to SARS-CoV-2, we used antibody titers against the ancestral SARS-CoV-2 (WA1) strain spike protein measured prior to the administration of booster doses, but predominantly after primary vaccination.

We interpreted the data in two ways. First, we estimated the probability of an individual having a PCR cycle threshold (Ct) value less than 30, as a proxy for infectiousness, on each day post detection using a logistic regression model. Second, we estimated the peak viral RNA concentrations, viral RNA proliferation rate, and viral RNA clearance rate across variants, immune statuses and age using a semi-mechanistic model. Our findings provide key estimates for the duration and magnitude of viral RNA shedding in the upper respiratory tract and its variation across symptom status, variants, immune states, and individuals.

### Data

We identified 2,875 distinct infections from 2,678 individuals in this cohort (**Supplementary Figure 1**). By the time of their final test, 2,460 (91.9%) individuals had one detected infection, 214 (7.99%) had two detected infections, three (0.11%) had three detected infections, and one (0.04%) had four detected infections. A total of 587 infections were detected within one day of a prior negative PCR test result, and thus the timing of the onset of test positivity can be assumed with reasonable accuracy. We defined these infections as the “frequent testing” group. The remaining 2,288 infections were detected two days or more from a previous negative test result or were detected with no prior negative test in the dataset. These were predominantly tests following suspected exposure, recent symptom onset, or periodic clearance for occupational health requirements, and thus we consider this latter group of detections as a reasonable proxy for infection detection in the absence of frequent testing, which is the case for most populations. We define these infections as the “delayed detection” group.

Of 1,086 infections with known symptom status, 766 reported symptoms at some point during the infection (70.5%). Individuals in the delayed detection group were more likely to be symptomatic than in the frequent testing group (73.1% vs. 64.9%; Chi-squared test statistic = 5.03; p-value <0.05). Most symptomatic individuals were detected around the time of symptom onset (**Supplementary Figure 2**; median delay from detection to symptom onset of zero days (N=553) in the delayed detection group and one day (N=171) in the frequent testing group). Symptom onset preceded the peak measured Ct value by a median of two days (N=550) in the delayed detection group and three days (N=171) in the frequent testing group (**Supplementary Figure 3**).

Based on genome sequencing, 1561 infections were confirmed to be Omicron (1 BA.2.10 isolate, the rest were lineages within BA.1), 266 confirmed to be Delta, and 247 confirmed as other lineages. An additional 801 infections were not sequenced; however, due to the rapid replacement of the circulating lineage in this cohort, we classified many of these as suspected Delta or other lineages based on the dominant lineage at time of detection (**Supplementary Figure 4**). We excluded non-sequenced samples following the detection of Omicron BA.1 due to the continued, albeit low-level, detection of Delta.

### Interpersonal variation in viral RNA trajectories

Viral trajectories varied substantially across individuals regardless of lineage (**Figure 1A**). Most individuals (65.5%, 95% CrI: 62.1-68.7%) in the frequent testing group remained potentially infectious, defined as having a Ct<30, on day 5 post detection. This fraction decreased to 20.0% (95% CrI: 17.3-22.9%) at day 10. In the delayed detection group, fewer individuals remained potentially infectious at days 5 and 10, likely because they were detected later in their infection. In this group, the probability of having a Ct<30 was 51.0% (95% credible interval (CrI): 48.2-53.6%) on day 5 post detection and 9.42% (95% CrI: 8.04-11.0%) on day 10.

**Figure 1.**
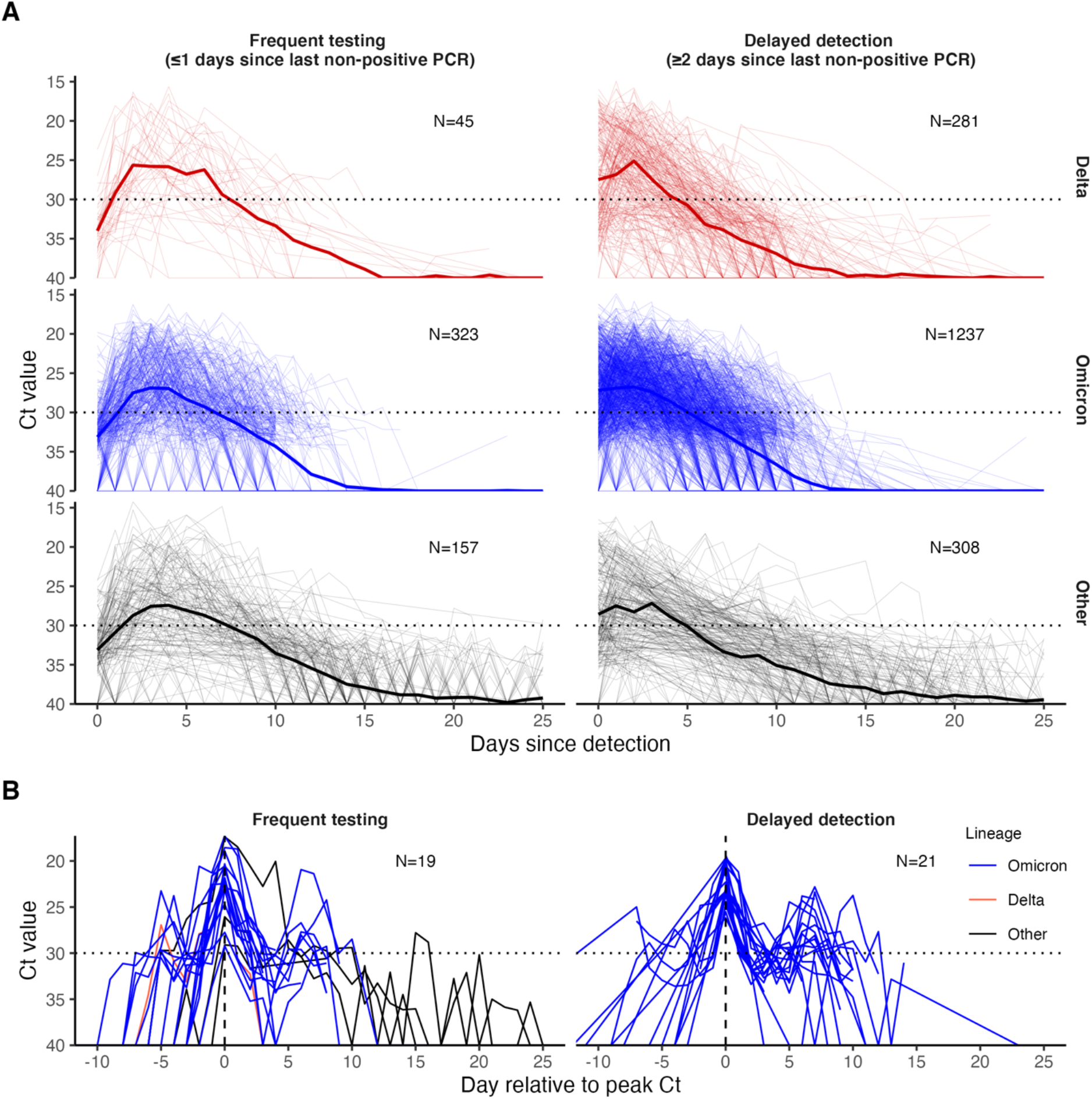
PCR Ct value trajectories for confirmed and suspected infections. **(A)** PCR Ct value trajectories for each acute Delta (red), Omicron BA.1 (blue), and other (black) infection. Individuals are grouped by the gap between detection and their most recent negative or inconclusive PCR test (Frequent testing vs. Delayed detection). Thick lines depict the mean Ct value over time, counting negative tests as Ct=40. Thin lines depict individual level Ct values over time. The horizontal dotted lines mark Ct=30, which we consider here as a proxy for possible infectiousness and antigen test positivity. (**B**) Subsets of PCR Ct value trajectories that were classified as rebounds, stratified by testing frequency group. Rebounds are defined here as any trajectory with an initial Ct value <30, followed by a sequence of two or more consecutive negative tests or tests with Ct value ≥30, and subsequently followed by two or more consecutive tests with Ct value <30.

### Incidence of rebounds

We next characterized the frequency of rebound viral RNA trajectories in this cohort. Viral rebounds may be characterized by the duration of the “quiescent” period of low viral concentration between distinct peaks, the duration of the subsequent rebound, and the timing of rebound onset relative to infection, but no consensus definition of viral rebound based on these quantities exists. We defined rebound as any viral trajectory with a decline in Ct value to <30 for 3+ days of tests (the rebound) after 3+ days of tests with Ct≥30 or a negative result (the quiescent period) following an initial Ct value <30 (the first detection of infection). Testing often ceased following initial clearance, and thus to minimize the impact of right censoring we only considered those trajectories with at least three days of tests with negative or Ct≥30 following a Ct value <30 as the denominator (N=999). We detected seven viral rebounds under this definition. Less stringent definitions led to more rebound classifications. For example, 40 (3.00%) of 1,334 infections were identified as rebounds when only 2+ consecutive days of Ct≥30 followed by 2+ days of Ct<30 was required to be classified as such (**Table 1**; **Figure 1B**). All individual-level viral trajectories classified as rebounds under this less stringent definition are shown in **Supplementary Figure S5**. Under this definition, we found that rebound infections were more likely in Omicron BA.1 infections, with 36 (4.10%; N=877) Omicron BA.1 infections resulting in rebound compared to one (0.562%; N=178) and three (1.08%; N=279) Delta and other infections, respectively (**Supplementary Table 1**; Chi-squared test for Omicron BA.1 (N=877) vs. non-Omicron BA.1 infection (N=457), test statistic = 9.69, p-value < 0.05). Similarly, we found that rebounds were more common in boosted individuals, with 32 (6.48%; N=494) rebounds in boosted individuals vs. three (0.929%; N=323) and two (1.26%; N=159) rebounds in vaccinated and unvaccinated individuals, respectively (**Supplementary Table 2**; Chi-squared test for boosted (N=494) vs. not-boosted (N=478) infection, test statistic = 18.1, p-value < 1e-4).

**Table 1.** Number of rebound infections classified under different definitions for initial clearance and subsequent rebound.

| Initial clearance duration (consecutive days with Ct≥30) | Rebound duration (days above Ct value threshold) | Ct value threshold of rebound | Rebounds | Total | Percentage |
| --- | --- | --- | --- | --- | --- |
| $\geq 4$ | $\geq 4$ | Ct<30 | 0 | 749 | 0.00% |
| $\geq 4$ | $\geq 3$ | Ct<30 | 1 | 749 | 0.13% |
| $\geq 4$ | $\geq 2$ | Ct<30 | 4 | 749 | 0.53% |
| $\geq 3$ | $\geq 4$ | Ct<30 | 2 | 999 | 0.20% |
| $\geq 3$ | $\geq 3$ | Ct<30 | 7 | 999 | 0.70% |
| $\geq 3$ | $\geq 2$ | Ct<30 | 16 | 999 | 1.60% |
| $\geq 2$ | $\geq 4$ | Ct<30 | 7 | 1334 | 0.53% |
| $\geq 2$ | $\geq 3$ | Ct<30 | 18 | 1334 | 1.35% |
| $\geq 2$ | $\geq 2$ | Ct<30 | 40 | 1334 | 3.00% |
| $\geq 4$ | $\geq 4$ | Ct<25 | 0 | 749 | 0.00% |
| $\geq 4$ | $\geq 3$ | Ct<25 | 0 | 749 | 0.00% |
| $\geq 4$ | $\geq 2$ | Ct<25 | 0 | 749 | 0.00% |
| $\geq 3$ | $\geq 4$ | Ct<25 | 1 | 999 | 0.10% |
| $\geq 3$ | $\geq 3$ | Ct<25 | 1 | 999 | 0.10% |
| $\geq 3$ | $\geq 2$ | Ct<25 | 2 | 999 | 0.20% |
| $\geq 2$ | $\geq 4$ | Ct<25 | 1 | 1334 | 0.08% |
| $\geq 2$ | $\geq 3$ | Ct<25 | 2 | 1334 | 0.15% |
| $\geq 2$ | $\geq 2$ | Ct<25 | 5 | 1334 | 0.38% |

### Minimal differences across variants and vaccination histories in the probability of having low Ct values over time

To assess differences in the duration of test positivity and infectiousness by variant and immune status, we modeled the probability of an individual having Ct<30 on each day post detection. As a baseline model, we fitted a logistic regression model with a smoothing spline on days since detection as a predictor. We compared various models with or without variant and vaccination history based on *k*-fold cross-validation and Bayesian model averaging.

The best-performing model used days since detection, stratified by both the cumulative number of previous exposures (infection or vaccination) and virus lineage, to predict the time course of low Ct values. The models including only vaccination status and lineage or days since previous exposure and lineage also performed well (**Supplementary Table 3 and 4**). This indicates that the interpersonal variation in low Ct values over time is better captured by models that account for exposure history than by models that account for time since detection alone. However, the difference in classification accuracy between the models was small. The baseline model, which included only the number of days since detection as a predictor, gave an overall classification accuracy for an individual having Ct<30 or ≥30/negative of 81.9% with an AUC of 88.7% (group-level classification accuracies: Ct<30 = 59.7%; Ct≥30/negative = 89.5%) for the frequent testing group, and an overall classification accuracy of 84.0% with an AUC of 90.4% (Ct<30 = 72.4%; Ct≥30/negative = 87.7%) in the delayed detection group. In contrast, the best model, which included the cumulative number of exposures and virus lineage in addition to days since detection, gave an overall classification accuracy of 82.8% with an AUC of 90.0% (Ct<30 = 63.7%; Ct≥30/negative = 89.4%) for the frequent testing group and an overall classification accuracy of 84.8% with an AUC of 91.2% (Ct<30 = 69.6%; Ct≥30/negative = 89.5%) in the delayed detection group. These results indicate that while exposure histories help to explain mean viral RNA kinetics, they provide little assistance in predicting an individual’s course of infectiousness over time, due to a high degree of individual-level variation, which may be dominated by stochastic effects or other unmeasured characteristics.

Vaccination provides multiple layers of protection against SARS-CoV-2, leading to reduced rates of infection (Tai et al., 2022) and faster clearance of the virus.(Kissler, Fauver, Mack, Tai, et al., 2021) Consistent with these findings, individuals who received two vaccine doses prior to infection with pre-Delta and pre-Omicron variants (N=17) cleared to negative results or high Ct values faster than unvaccinated individuals (N=216) (**Supplementary Figure 6**). While boosting reduced rates of infection in our cohort,(Tai et al., 2022) boosted individuals with Omicron BA.1 infections (N=651) tended to sustain low Ct values for longer durations than individuals who had only undergone an initial vaccine course (N=251), defined as either two doses of an mRNA vaccine or a single dose of the Ad.26.COV2.S adenovirus vector-based vaccine (**Figure 2A; Supplementary Figure 6**). This pattern was robust to refitting the model after excluding player infections, resulting in a subpopulation more representative of the general population in age and health status (**Supplementary Figure 7**). We also found similar patterns after subsetting infections by their symptom status (**Supplementary Figure 8**).

**Figure 2.**
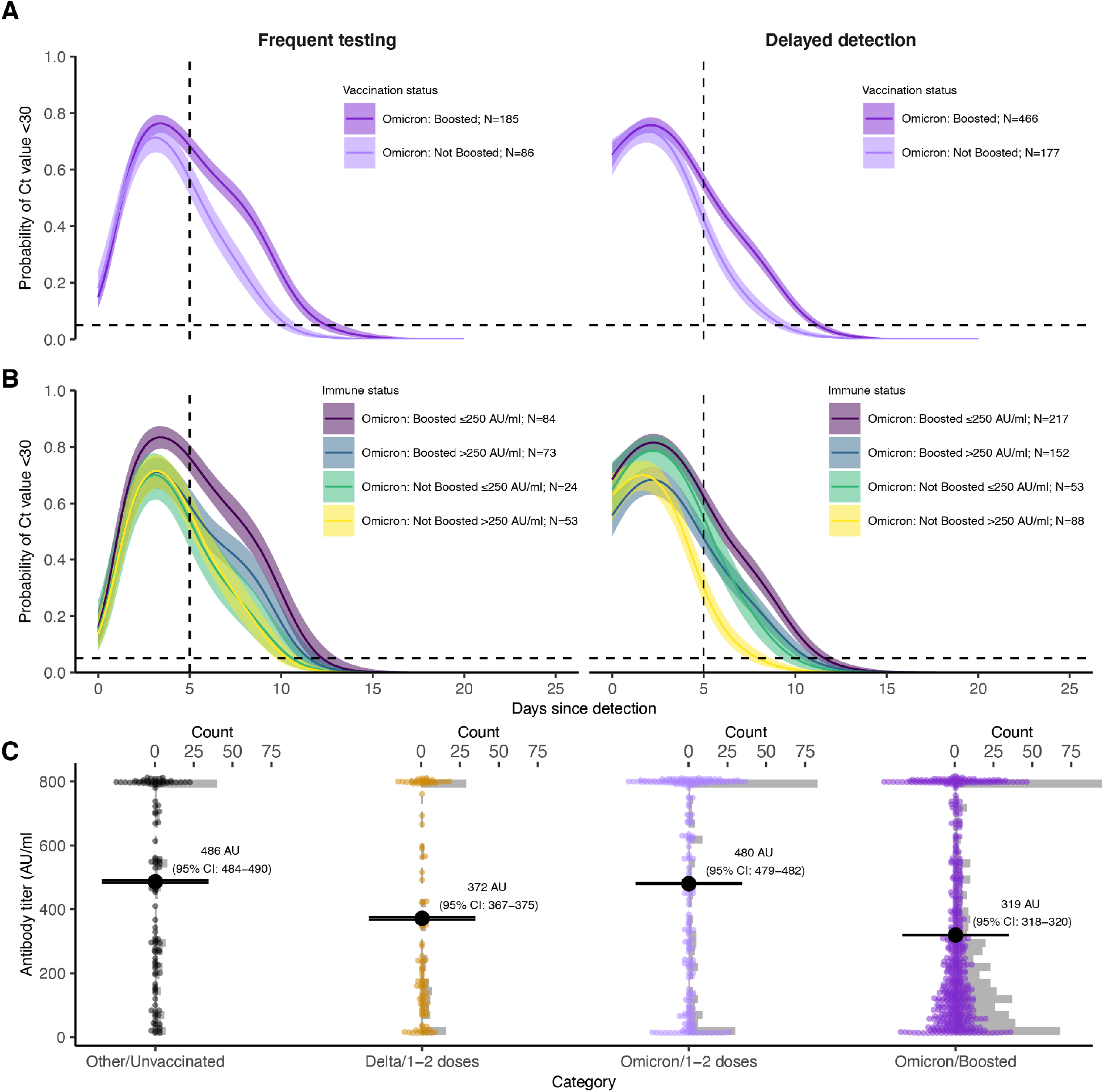
(**A**) Proportion of confirmed and suspected Omicron BA.1 infections with Ct<30 on each day post detection by vaccination status and detection group. Solid colored lines and shaded ribbons depict posterior estimates from a generalized linear model predicting probability of Ct<30 as a function of days since detection, vaccination status and variant, showing the posterior mean (solid line) and 95% credible intervals (shaded ribbon) of each conditional effect. Dotted horizontal and vertical lines mark 5% probability and day 5 post detection, respectively. (**B**) As in (A), but additionally stratified by single point-in-time anti-spike antibody titer against the ancestral SARS-CoV-2 (WA1) strain as measured by the Diasorin Trimeric Assay. (**C**) Distribution of antibody titers (colored points) stratified by variant and vaccination status of each detected infection, with mean titers (large black points) and bootstrapped 95% confidence intervals for the mean (horizontal lines). Grey bars are histograms of antibody titer counts in bins of 10 arbitrary units (AU)/ml. Note that stratification is by infection and not individual, and that antibody titers were measured at a single point in time rather than near the time of infection. The Diasorin Trimeric Assay values are truncated between 13 and 800 AU/ml.

We also considered a possible effect of age, as boosted individuals in this cohort were typically older than non-boosted individuals at the time of BA.1 infection (mean age of 37.6 years in the BA-1 infected, boosted group vs. 31.3 years in the BA.1-infected, non-boosted group), and thus the differences between vaccination groups could be driven by an age-specific effect. Refitting the regression model including an additional spline term for the interaction of days since detection and age group (categorized as <30, 30-50 or >50 years old) suggested that older individuals did maintain Ct<30 for longer on average than younger individuals after conditioning on vaccination status (**Supplementary Figure 9A)**. However, the effect of a higher proportion with Ct<30 in boosted individuals relative to non-boosted individuals also remained after conditioning on age, suggesting that both older age and booster status explain some variation in duration of Ct<30 (**Supplementary Figure S9B**). Furthermore, models including age were universally better supported in the model comparison analysis and provided an improvement in classification accuracy, but in both cases the gains were small.

### Pre-Omicron antibody titer explains variation in viral RNA clearance

To assess the mechanisms behind the unexpected slower clearance in boosted Omicron BA.1 infections, we assessed viral kinetics stratified by antibody titer. In addition to exposure history information, 979 individuals were tested at least once (1,017 measurements total) with the Diasorin Trimeric Assay for antibody titers against the spike protein from the ancestral SARS-CoV-2 (WA1) strain (**Supplementary Figure 10**). Most titers were obtained from mid-September to mid-October 2021, and thus we consider these titers to represent an individual’s post primary vaccination course response rather than post-boost/post-Omicron infection immunity (**Supplementary Figure 11**). The median time between the most recent vaccine dose and the titer draw was 162 days (interquartile range: 129–180 days) (**Supplementary Figure 12**).

We hypothesized that these single point-in-time SARS-CoV-2 antibody titer measurements represented a proxy of the strength of the immune response to SARS-CoV-2 and thus would be reflected in the features of viral kinetics over the course of infection. A total of 494 measurements were classified as low antibody titers (≤250 arbitrary units [AU]/ml) and 523 as high titers (>250 AU/ml). This cutoff was chosen as a conservative upper bound for defining risk of Delta infection (see **Materials & Methods**). Most individuals either had very low titers (<13 AU/ml) or titers at the upper limit of detection (800 AU/ml) of the assay (**Figure 2C)**.

We fitted a logistic regression model for the probability of having a Ct<30 on each day since detection, stratified by the interaction of an individual’s booster status and their pre-booster antibody titer status (**Figure 2B; Supplementary Figure 13**). Boosted individuals with a low antibody titer had the highest and longest duration of Ct<30 over time since detection in both the frequent testing and delayed detection group. In the delayed detection group, individuals with low antibody titers were more likely to have Ct<30 than individuals with high antibody titers regardless of booster status, though boosted individuals with high antibody titers maintained Ct<30 for longer than non-boosted individuals with low titers. To account for potential confounding from waning immunity, in which low titers simply represent a longer time since previous exposure, we restricted the dataset to include only individuals who had their titer measured within 100-200 days after a previous exposure. We also repeated the analysis after restricting to only infections detected 60-90 days following an antibody titer measurement. These trends were maintained in both sensitivity analyses (**Supplementary Figure 14**).

Based on these findings, we hypothesized that boosted individuals who nevertheless were infected with Omicron BA.1 may have had relatively poor BA.1-specific immune responses to prior SARS-CoV-2 exposures, leading to uncharacteristically long infections. To test this hypothesis, we stratified each infection by vaccination status and variant and compared antibody titers between these groups (**Figure 2C**). Antibody titers were lower among fully vaccinated individuals who were subsequently infected with Delta than individuals who had been infected with a pre-Delta variant. This suggests that individuals with a high antibody titer at around the time of Delta circulation were less likely to be infected with Delta. In contrast, we found that mean antibody titers among Omicron BA.1-infected, fully vaccinated individuals were similar to individuals in the pre-Delta, unvaccinated group, suggesting that higher titer individuals were not substantially less likely to be infected than lower titer individuals. Finally, we found that antibody titers were lowest among Omicron BA.1 infected boosted individuals, suggesting that individuals with a high titer measurement prior to being boosted were less likely to have Omicron BA.1 infections.

These results are consistent with an additional age group-level effect also contributing towards differences in the proportion of individuals with Ct<30 over time. We found that younger BA.1-infected individuals had higher antibody titers on average than older BA.1-infected individuals, but that still BA.1-infected boosted individuals had consistently lower mean antibody titers than BA.1-infected non-boosted individuals within each age group (**Supplementary Figure S15A**). Furthermore, refitting the logistic regression model including terms for both age group and the interaction of vaccination status and antibody titer group showed a consistent trend of boosted, low titer individuals maintaining Ct<30 for longer than other high titer individuals within each age group (**Supplementary Figure S15B**). However, we note that at this level of stratification, the sample sizes for each subgroup are small and thus there is considerable uncertainty for some combinations of age group, titer group and vaccination status.

### The effect of immune status and variant on viral proliferation, peak viral RNA titers, and clearance

We next adapted a framework to estimate the impact of antibody titer, vaccination status, and variant on peak viral RNA concentrations, proliferation phase duration, and clearance duration (**Figure 3**).(Kissler, Fauver, Mack, Olesen, et al., 2021; Kissler, Fauver, Mack, Tai, et al., 2021) According to the viral kinetic model, and among Omicron BA.1 infections, boosted individuals had a longer estimated viral clearance time than non-boosted individuals (8.4 days (95% CrI: 8.0-8.7) *vs*. 6.2 days (95% CrI: 5.8-6.6), respectively), in line with the results from the logistic regression model. Viral proliferation times and peak viral RNA were similar among boosted and non-boosted individuals with Omicron BA.1 infections (**Supplementary Table 5**). When stratifying by post-initial vaccination antibody titer, Delta infections featured a consistently higher peak viral RNA than Omicron BA.1 infections. Among Omicron BA.1 infections, high antibody titers were associated with faster viral clearance times and lower peak viral RNA. Proliferation times were similar across variants and titers (**Supplementary Table 6**).

**Figure 3.**
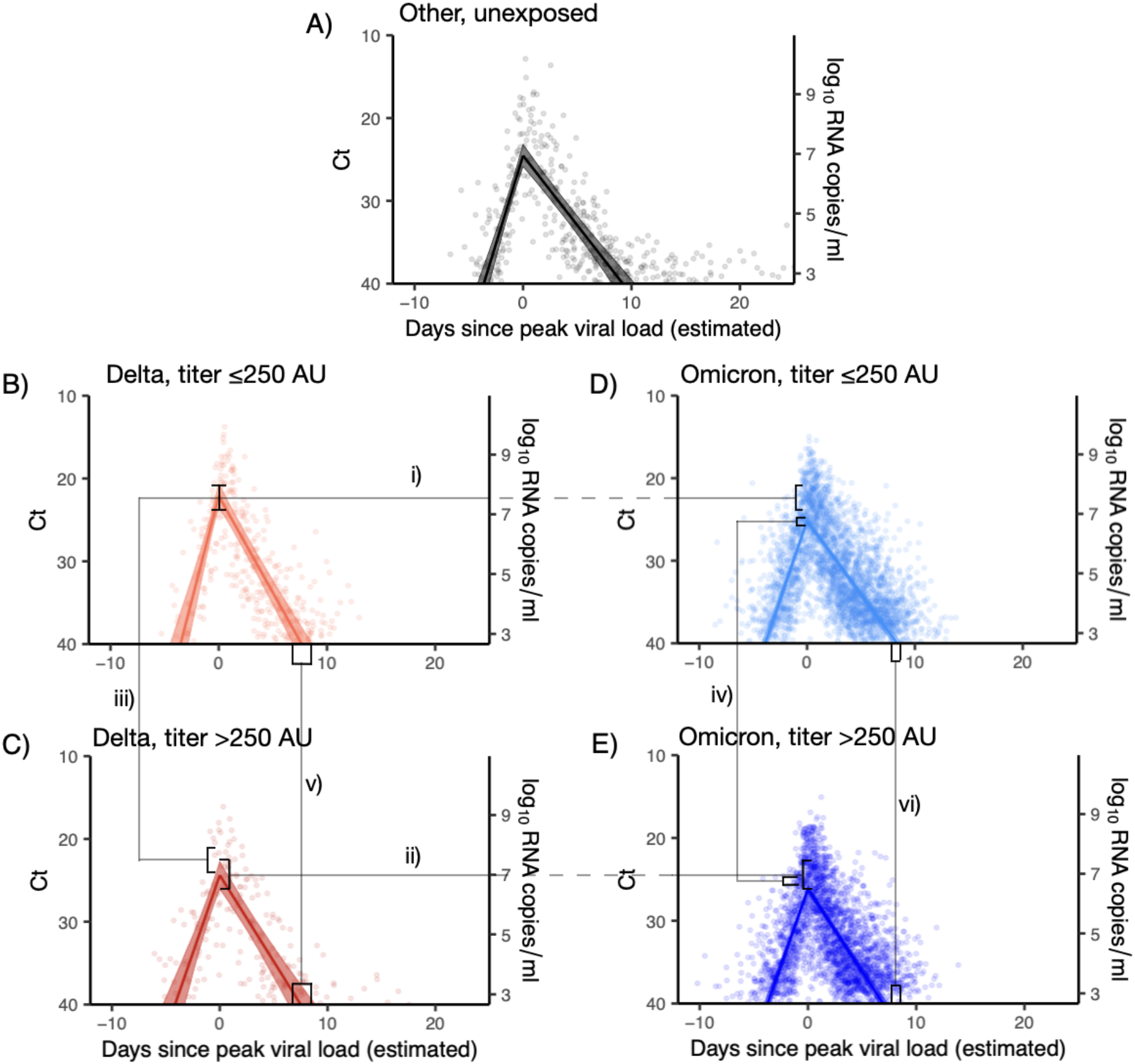
Estimated viral trajectories by variant and titer. Points depict measured Ct values, lines depict the estimated population mean viral trajectories, and shaded regions depict the 95% credible intervals for the estimated population viral trajectories. A) Non-Delta and non-Omicron infections in individuals who were previously unexposed (no prior record of vaccination or infection), B) Delta infections with titer ≤ 250, C) Delta infections with titer > 250, D) Omicron infections with titer ≤ 250, E) Omicron infections with titer > 250. Peak viral loads were higher for Delta infections than for Omicron infections when stratifying by titer (i and ii), and titers ≤ 250 were associated with higher viral loads when stratifying by variant (iii and iv). Low titers were also associated with longer clearance times (v and vi).

We fitted the viral RNA kinetic model to Omicron BA.1 infections after stratifying individuals based on their symptom status as well as vaccination status or antibody titer group. We found the same pattern of longer clearance times for boosted individuals relative to fully vaccinated individuals, with symptomatic boosted individuals demonstrating longer clearance times than asymptomatic boosted individuals (**Supplementary Table 7**). Among those with low antibody titer, presence of symptoms was associated with higher peak viral RNA and longer clearance times, while for those with high antibody titer, peak viral RNA and clearance times were similar between symptom statuses (**Supplementary Table 8**).

Finally, we fitted the models allowing for BA.1 viral RNA kinetics to vary by age in addition to vaccination status or antibody titer. Consistent with the logistic regression results, older individuals demonstrated longer average clearance times than younger individuals across vaccination and antibody titer groups. When stratifying by immune status, individuals aged 50+ years took between roughly 1–2 days longer to clear than individuals aged 30–50, and 2–3 days longer to clear than individuals under 30 (**Supplementary Tables 9 & 10**). However, we still found a consistent effect of antibody titer and booster status on clearance time despite this additional age effect. Individuals with low antibody titer and boosted individuals took 1-2 days longer to clear than individuals with high antibody titer and non-boosted individuals in both the <30- and 30–50-year age groups (**Supplementary Tables 9 & 10**).

## Discussion

We found that individuals infected with SARS-CoV-2 often had Ct values <30 beyond the five-day isolation period following SARS-CoV-2 infection currently recommended by the CDC.(Centers for Disease Control and Prevention, n.d.) This finding is in line with other studies measuring Ct values from upper respiratory tract samples, the duration of antigen test positivity, and the duration of infectious viral load or culturable virus.(Boucau, Marino, et al., 2022; Earnest et al., 2022; Ke et al., 2022; Landon et al., 2022; Lefferts et al., 2022) While we do not have data on infectiousness by day to clarify the exact link between Ct and infectiousness, nearly half of the individuals in this cohort had potentially infectious viral loads (Ct<30) five days after their initial detection, even in those detected later in their infection course (Singanayagam et al., 2020a). By day 10, the number of individuals with Ct<30 was substantially reduced but still high. The duration of positivity was highly variable across individuals, and low Ct values consistent with potential infectiousness were sometimes maintained for up to two weeks. These observations suggest the use of test-based, rather than time-based, protocols for defining the duration of isolation to limit the spread of SARS-CoV-2.

Rebounds with recurrence of symptoms and positive rapid antigen tests after a period of negative test results have been increasingly reported in individuals treated with SARS-CoV-2 antiviral drugs,(Boucau, Uddin, et al., 2022; Charness et al., 2022) but estimates for the frequency of viral rebounds in the absence of antiviral treatment have been lacking. Among infected boosted individuals in this cohort, who were predominantly infected with Omicron BA.1, we detected seven rebounds in viral trajectory, stringently defined as any Ct value trajectory with at least 3 consecutive days of negative tests or tests with Ct≥30 after the initial peak followed by 3 or more consecutive days with Ct<30. However, more rebounds were detected when using less stringent Ct value-based definitions and were more frequent in Omicron-infected or boosted individuals, occurring in ∼6% of infections in contrast to ∼1% of infections in the pre-booster pre-Omicron phase of the pandemic. It was not routine for testing to continue following suspected clearance in this cohort, and thus these results may represent a lower bound on the incidence of rebound infections. The frequency of viral trajectory rebounds depends on the definition of ‘rebound’, highlighting the need for standardized definitions to enable comparisons across studies. We did not measure the recurrence of culturable virus during these resurgent low Ct periods, and thus further work is needed to understand if viral RNA rebounds are a reliable proxy for infectivity. Moreover, we did not have sufficient information to define rebounds with respect to clearance and recurrence of symptoms, though the experience of the occupational health team is that rebounds of a clinical nature have been extremely rare, with only one documented case.(Mack et al., 2021) Overall, these findings suggest that symptom monitoring after clearing isolation may be warranted, and a return to isolation may be necessary for individuals with rebound infections.(Charness et al., 2022)

Boosted individuals in this cohort were less likely to be infected with Omicron BA.1,(Tai et al., 2022) and those who had a breakthrough infection tended to have a low antibody titer measurement to the WA1 spike protein after their initial vaccine course. In this context, test positivity following Omicron BA.1 infection lasted longer for boosted individuals than for non-boosted individuals, regardless of symptom status. This observation was further supported by a viral kinetic model that found longer clearance times for Omicron BA.1 infections in boosted relative to non-boosted individuals. Moreover, high antibody titers to the WA1 spike protein were associated with lower peak viral RNA concentrations and faster clearance times for both Delta and Omicron BA.1 infections. Together, these results suggest that the low antibody titers in infected boosted individuals conferred increased risk for infection as well as slower control and clearance of infection.

The effect of age on viral kinetics complicates the interpretation of these findings. Prior to the detection of Omicron BA.1, older individuals have been found to take longer to clear infection on average than younger individuals.(Caputo et al., 2021; Cevik et al., 2021; Jones et al., 2021; Long et al., 2021; Néant et al., 2021; Singanayagam et al., 2022) However, these findings are not unequivocal, as a previous systematic review found the effect of age on viral kinetics was diminished after accounting for disease severity.(Chen et al., 2021) Our data support an effect of age on viral clearance times, with longer times from peak to clearance in individuals >50 years compared to those <30 years regardless of lineage and immune state. In this cohort, older individuals were more likely to be boosted prior to becoming infected with BA.1 than younger individuals, and thus the finding of delayed clearance in BA.1-infected, boosted individuals can be partially attributed to delayed clearance in older individuals. However, we found consistent delayed clearance in boosted relative to non-boosted individuals within each age group, notably in the <30 years group. Furthermore, the pattern of lower antibody titers to WA1 spike in BA.1-infected, boosted individuals relative to BA.1-infected, non-boosted individuals was also consistent within each age group, suggesting that low WA1 spike titers correlate with increased infection risk and slower clearance in addition to any age-specific effects.

An important limitation of this study is that the cohort is not representative of the general population, as it is predominately male, young, and includes professional athletes. However, our key findings were preserved in analyses after excluding the players. We did not test for the presence of infectious virus, and our findings are based on Ct values obtained from combined nasal and oropharyngeal swabs.(Ke et al., 2022) While low Ct values have been associated with potential infectiousness and antigen test positivity,(Bullard et al., 2020; Jaafar et al., 2021; Jefferson et al., 2020; Singanayagam et al., 2020b) this is an imperfect proxy. It is possible that some infections were undetected, and thus the reported number of prior infections should be interpreted as a lower bound for each member of the cohort. SARS-CoV-2 antibody titers were only measured from mid-September to mid-October 2021 and were taken at varying time points after initial vaccination course (between 0 and 290 days), so we could not assess the relationship between antibody waning and viral kinetics. Antibody titers were measured against the spike protein of the WA1 lineage, which correlate poorly with protection against the antigenically distinct Omicron lineages; thus, it is unclear how these data are associated specifically to Omicron-immunity, beyond representing a proxy for overall immune response.

Variants and immune statuses interact, sometimes in unexpected ways, to produce viral kinetics that differ in duration and intensity. Collecting longitudinal viral load data in more diverse cohorts will help to ensure that isolation and quarantine policies are based on the best available evidence and will help to properly contextualize results from ongoing drug and vaccine trials. Similarly, our findings suggest that SARS-CoV-2 control measures may be better informed by measurements of immune status than proxies such as number or timing of receipt of vaccine doses or of infections. Testing this hypothesis will require widespread collection and analysis of serological, infection, and vaccination data in diverse cohorts and broader availability of quantitative antibody tests designed for the spike protein of Omicron lineages.

## Methods

### Study design

The data reported here represent a convenience sample including team staff, players, arena staff, vendors, and others affiliated with the NBA as described previously.(Kissler, Fauver, Mack, Olesen, et al., 2021; Kissler, Fauver, Mack, Tai, et al., 2021) The retrospective study includes samples collected between 7th July 2020 and 26th January 2022 (**Supplementary Figure 1**). Clinical samples were obtained by combined swabs of the anterior nares and oropharynx, collected separately from each anatomical site, for each patient administered by a trained provider. Daily testing was required for most individuals prior to vaccination availability, with less frequent testing but close monitoring required after vaccination. Cycle threshold (Ct) values were generated using the Roche cobas target 1 assay. For the viral kinetics model analyses, Ct values were converted to viral genome equivalents using a standard curve.(Kissler, Fauver, Mack, Olesen, et al., 2021)

We classified all individuals as having Ct value <30 or not on each day post-detection. This threshold was chosen based on a combination of antigen sensitivity and studies of virus culture by Ct, where the presence of culturable virus is often assumed to correlate with infectivity.(Brihn et al., 2021; Bullard et al., 2020; Pilarowski et al., 2021; Singanayagam et al., 2020b; Thommes et al., 2021) We stratified infections by those who had a negative or inconclusive test ≤1 day prior to detection and those whose last negative or inconclusive test was ≥2 days ago. We assumed that individuals testing negative at the end of an acute infection remained negative for the remainder of the study period, whereas those ending in a positive test are right-censored. Rebound trajectories were defined as any trajectory with a sequence of two or more consecutive Ct values ≥30 or negative tests after the initial peak followed by two or more consecutive Ct values <30. We considered more stringent definitions both for initial clearance (3+ or 4+ days of Ct ≥30 or negative test following initial peak) and subsequent rebound (3+ or 4+ days of Ct < 30). In some instances, individuals were tested multiple times per day and thus for ease of model fitting we excluded 3,751 positive or inconclusive and 14,713 negative samples from repeat tests on the same day in our analyses, prioritizing the earliest test and then lowest Ct value test on each day.

Vaccination information was reported and verified by NBA staff and a clinical operational team. 828 individuals had been boosted by the time of their last PCR test, 529 had completed their primary vaccination course (two doses of an mRNA vaccine or one dose of Janssen / Ad.26.COV2.S adenovirus vector-based vaccine), 8 had received one vaccine dose, and 13 confirmed to be unvaccinated. The vaccination statuses of the remaining individuals were unknown. The time course of individual vaccination and exposure times is shown in **Supplementary Figure 11**.

### Study oversight

In accordance with the guidelines of the Yale Human Investigations Committee, this work with de-identified samples was approved for research not involving human subjects by the Yale Institutional Review Board (HIC protocol # 2000028599). This project was designated exempt by the Harvard Institutional Review Board (IRB20-1407).

### Classification of infections

We tagged each series of positive tests buffered by at least 14 days of negative or missing tests on each side as a distinct infection. After an infection was flagged, subsequent positives were not classified as a new infection for 90 days. Isolated positive tests with no other positive within 14 days either side were not considered as detections. We track the cumulative number of exposures (defined as either receiving a vaccination or infection) over time. Individuals who received the Janssen/Ad.26.COV2.S adenovirus vector-based vaccine were counted as having received two vaccine doses. 351 additional infections were reported to the program outside of the main testing regime, either through an external PCR or rapid antigen test, or from a positive antibody test result (not including the Diasorin Trimeric Assay results described below). We consider these detections as contributing towards an individual’s infection history but are unable to include them in the Ct value trajectory analyses.

### Genome sequencing and lineage assignment

RNA was extracted and confirmed as SARS-CoV-2 positive by RT-qPCR.(Vogels et al., 2021) Next Generation Sequencing was performed with the Illumina COVIDSeq ARTIC viral amplification primer set (V4, 384 samples, cat# 20065135). Library preparation was performed using the amplicon-based Illumina COVIDseq Test v033 and sequenced 2×74 on Illumina NextSeq 550 following the protocol as described in Illumina’s documentation. The resulting FASTQs were processed and analyzed on Illumina BaseSpace Labs using the Illumina DRAGEN COVID Lineage Application;(*BaseSpace Labs. DRAGEN COVID Lineage*, 2021) versions included are 3.5.0, 3.5.1, 3.5.2, 3.5.3 and 3.5.4. The DRAGEN COVID Lineage pipeline was run with default parameters recommended by Illumina. Lineage assignment and phylogenetics analysis using the most updated version of Pangolin(Rambaut et al., 2020) and NextClade(Aksamentov & Neher, 2021), respectively. All sequenced Omicron infections were lineage BA.1 apart from 1 BA.2.10 infection. Sequenced Delta infections were a combination of lineages B.1.617.2 and AY.x.

There were 3 and 482 non-sequenced infections in the window of time when Alpha was replaced by Delta (29^th^ May 2021 to 18^th^ July 2021) and after the first detection of Omicron BA.1 (3^rd^ December 2021 onwards), respectively (**Supplementary Figure 4**). We removed these 485 infections from variant-specific analyses and assigned all non-sequenced infections prior to the detection of Omicron BA.1 to the dominant lineage at the time of detection (i.e., all infections prior to 29^th^ May 2021 were assumed “Other” and all infections between 18^th^ July 2021 and 3^rd^ December 2021 were assumed “Delta”). We removed all non-sequenced infections detected after 3^rd^ December 2021 from variant-specific analyses rather than classifying them as Omicron BA.1 due to the continued presence of Delta. Omicron BA.2 was not regularly detected until after this period, with only 1 confirmed BA.2 infection (BA.2.10), which was removed from these analyses.

### Antibody titers

Individuals were tested with the Diasorin Trimeric Assay for IgG antibody titers against the ancestral SARS-CoV-2 (WA1) strain spike protein during the 2021 pre-season period (September-October 2021). The majority (>90%) of blood draws were from mid-September to early October 2021. We classified individuals with a titer of >250 AU/ml as being in the high titer group and in the low titer group otherwise, chosen based on its correlation with authentic virus neutralization results for wildtype and Delta.(Liu et al., 2021; Wang et al., 2021) Specifically, an authentic virus neutralization titer of 100 was found to be well correlated with a 50% protective neutralization level for wildtype(Khoury et al., 2021) and found to correspond to a DiaSorin AU of 189.09 (95%CI: 147.61-235.75) (**Supplementary Figure 16**). The cutoff of 250 was therefore chosen as a conservative upper bound classifying an individual as at lower risk of infection with Delta or wildtype SARS-CoV-2. Note that this cutoff does not predict infection risk with Omicron and was simply chosen as a proxy for an individual’s immune competence.

### Logistic regression models

We used the RStan package *brms* to fit Bayesian logistic regression models estimating the probability of having Ct value <30 on each day post detection, stratified by detection speed.(Bürkner, n.d.) As a baseline, we considered a model without variant-specific effects, using smoothing splines to estimate the probability of having a Ct value less than 30 on each day post detection. We then fitted additional logistic regression models, adding additional spline terms capturing the effect of vaccination status, cumulative number of previous exposures or days since previous exposure, and/or lineage with days since detection. We then ranked models based on the expected log predictive density calculated using *k*-folds cross-validation (25 folds) and evaluated their classification accuracy and area under the receiver operator curve. For the antibody titer analyses, we fitted Bayesian logistic regression models for the probability of Ct value <30 as a function of days since detection, stratified by the interaction of titer group (above or below 250 AU/ml), lineage and vaccination status. To account for possible confounding from age effects, we ran sensitivity analyses refitting all of the logistic regression models including an additional spline term for the effect of age group on probability of Ct value <30 on each day since detection. We grouped individuals into three age groups: <30 years, 30-50 years, >50 years. Further details on the fitting process can be found in the **Supplementary Methods**.

### Viral kinetic model

We extended a previously reported model for capturing SARS-CoV-2 viral kinetics to estimate the viral proliferation time, viral clearance time, and peak viral load by variant and immune status.(Kissler, Fauver, Mack, Olesen, et al., 2021; Kissler, Fauver, Mack, Tai, et al., 2021) The model approximates viral kinetics on a logarithmic scale as a piecewise linear function, corresponding to an exponential increase of virus followed by an exponential clearance at possibly different rates. To estimate the relationship between booster status and viral kinetics, we first stratified the model by (1) Omicron boosted and (2) Omicron non-boosted individuals. There were too few boosted individuals who were infected with other variants to reliably fit the model to non-Omicron infections. Next, to estimate the relationship between antibody titer and viral kinetics, we stratified the model by (1) Delta infections with titer ≤ 250, (2) Delta infections with titer > 250, (3) Omicron infections with titer ≤ 250 (4) Omicron infections with titer > 250, and finally (5) non-Delta and non-Omicron infections in individuals who had not had any prior exposure either through infection or vaccination, to serve as a baseline. Full details on the fitting procedure may be found in (Kissler, Fauver, Mack, Olesen, et al., 2021; Kissler, Fauver, Mack, Tai, et al., 2021) and the **Supplementary Methods**.

## Data Availability

Code and data to regenerate these analyses, including individual cycle threshold value trajectories, are available at https://github.com/gradlab/SC2-kinetics-immune-history.

