## Supplementary Material for "Quantifying the impact of immune history and variant on SARS-CoV-2 viral kinetics and infection rebound: a retrospective cohort study"

### Supplementary methods

#### Logistic regression models

Model fitting.

We fitted Bayesian logistic regression models for the probability of an individual having Ct value <30 on each day post detection using the *brms* package version 2.14.4. Models were run on the Harvard FAS Research Computing cluster using R version 4.0.2. For each model, we ran 4 chains for 2000 iterations each. Weakly informative priors (normal distributions with means of 0 and standard deviations of 10) were used for all model parameters. We assessed convergence based on all estimated parameters having a Gelman R-hat statistic less than 1.1.

#### Viral kinetics model

Statistical analysis.

Following previously described methods,[1,2] we used a Bayesian hierarchical model to estimate the proliferation duration, clearance duration, and peak viral concentration for acute SARS-CoV-2 infections, stratified by variant (Omicron, Delta, Other),immune status (vaccination history, including unexposed, 1-2 doses, or boosted; and antibody titer, including unexposed, titer ≤ 250 AU, and titer > 250 AU) and age (groups of <30, 30-50, and >50 years old). The model describes the $\log_{10}$ viral concentration during an acute infection using a continuous piecewise-linear curve with control points that specify the time of acute infection onset, the time and magnitude of peak viral concentration, and the time of acute infection clearance. The assumption of piecewise linearity is equivalent to assuming exponential viral growth during the proliferation period followed by exponential viral decay during the clearance period. The control points were inferred using the Hamiltonian Monte Carlo algorithm as implemented in Stan (version 2.24).[3] We used priors informed by a previous analyses.[1,2] Data and code are available online.

Model fitting.

To restrict to a set of well-observed acute infections for model fitting, we first removed any sequences of 3 or more consecutive negative tests (Ct = 40) from each acute infection to avoid overfitting to these trivial values. We kept only acute infections with at least one Ct value < 32 and at least 3 Ct values < 40 (the limit of detection).

We constructed a piecewise-linear regression model to estimate the peak Ct value, the time from infection onset to peak (*i.e.* the duration of the proliferation stage), and the time from peak to infection resolution (*i.e.* the duration of the clearance stage). This is represented by the equation

$$E[Ct(t)] = \begin{cases} \text{l.o.d} - \frac{\delta}{t_p - t_o}(t - t_o) & t \leq t_p \\ \text{l.o.d} - \delta + \frac{\delta}{t_r - t_p}(t - t_p) & t > t_p \end{cases}$$

Here, E[*Ct(t)*] represents the expected value of the Ct at time *t,* "l.o.d" represents the RT-qPCR limit of detection, $\delta$ is the absolute difference in Ct between the limit of detection and the peak (lowest) Ct, and $t_o$, $t_p$, and $t_r$ are the onset, peak, and recovery times, respectively.

Before fitting, we re-parametrized the model using the following definitions:

- $\Delta Ct(t)$ = l.o.d. – $Ct(t)$ is the difference between the limit of detection and the observed Ct value at time *t.*
- $\omega_p = t_p - t_o$ is the duration of the proliferation stage.

- $\omega_r = t_r - t_p$ is the duration of the clearance stage.

We next characterized the likelihood of observing a given $\Delta Ct(t)$ using the following mixture model:

$$L(\Delta Ct(t) = x|\delta, t_p, \omega_p, \omega_r) = (1-\lambda)\Big[f_N(x|E[\Delta Ct(t)], \sigma(t)) + I_{lod} F_N(0|E[\Delta Ct(t)], \sigma(t))\Big] + \lambda f_{\mathrm{Exp}}(x|\kappa)$$

The left-hand side of the equation denotes the likelihood ($L$) that the observed viral load, as measured by Ct deviation from the limit of detection ($\Delta Ct(t)$), is equal to some quantity $x$ given the model parameters $\delta$ (peak viral load), $t_p$ (time of peak viral load), $\omega_p$ (proliferation time), and $\omega_r$ (clearance time). This likelihood is equal to the sum of two main components: the likelihood that the observed value was generated by the modeled viral kinetic process, denoted by the bracketed term preceded by a ($1-\lambda$); and the likelihood that the observed value was a false negative, denoted by the term preceded by a $\lambda$. In the bracketed term representing the modeled viral kinetic process, $f_N(x \mid E[\Delta Ct(t)], \sigma(t))$ represents the Normal PDF evaluated at $x$ with mean $E[\Delta Ct(t)]$ (generated by the model equations above) and observation noise $\sigma(t)$. $F_N(0 \mid E[\Delta Ct(t)], \sigma(t))$ is the Normal CDF evaluated at 0 with the same mean and standard deviation. This represents the scenario where the true viral load goes below the limit of detection, so that the observation sits at the limit of detection. $I_{lod}$ is an indicator function that is 1 if $\Delta Ct(t) = 0$ and 0 otherwise; this way, the $F_N$ term acts as a point mass concentrated at $\Delta Ct(t) = 0$. Last, $f_{\mathrm{Exp}}(x \mid \kappa)$ is the Exponential PDF evaluated at $x$ with rate $\kappa$. We set $\kappa = \log(10)$ so that 90% of the mass of the distribution sat below 1 Ct unit and 99% of the distribution sat below 2 Ct units, ensuring that the distribution captures values distributed at or near the limit of detection. We did not estimate values for $\lambda$ or the exponential rate because they were not of interest in this study; we simply needed to include them to account for some small probability mass that persisted near the limit of detection to allow for the possibility of false negatives. A schematic of the likelihood function is depicted in **Supplementary Figure 17.**

We used a hierarchical structure with a non-centered parameterization to describe the distributions of $\omega_p$, $\omega_r$, and $\delta$ for each person:

$\omega_p[i] = \mathrm{Exp}[\mu_{\omega p} + \zeta^i_{wp} + \sigma_{wp} N^i_{wp}]\ \omega^*_p$
$\omega_r[i] = \mathrm{Exp}[\mu_{\omega r} + \zeta^i_{wr} + \sigma_{wr} N^i_{wr}]\ \omega^*_r$
$\delta[i] = \mathrm{Exp}[\mu_\delta + \zeta^i_\delta + \sigma_\delta N^i_\delta]\ \delta^*$

Here, $\omega^*_p$, $\omega^*_r$, and $\delta^*$ are user-defined estimated values for the means of $\omega_p$, $\omega_r$, and $\delta$, so that the exponential terms represent an adjustment factor relative to that midpoint defined by $\mu$ (a shared adjustment factor for the entire population), $\zeta^i$ (an adjustment factor shared among individuals of a given variant/immune category), and $\sigma$ (a shared standard deviation for the entire population). The $N^i$ terms represent individual-level random effects. The prior distributions for the $\mu$, $\zeta$, and $\sigma$ terms were all Normal(0, 0.25) (with $\sigma$ truncated to have support on the positive reals). These prior distributions define LogNormal adjustment factors that have ~99% of their probability mass between 0.5 and 2, so that the prior distributions for $\omega_p$, $\omega_r$, and $\delta$ cover roughly half to twice their prior estimated midpoint values.

We used a Hamiltonian Monte Carlo fitting procedure implemented in Stan (version 2.24) and R (version 3.6.2) to estimate the parameters. We ran four MCMC chains for 2,000 iterations each with a target average proposal acceptance probability of 0.8. The first half of each chain was discarded as the warm-up. The Gelman R-hat statistic was less than 1.1 for all parameters. This indicated good overall mixing of the chains. There were no divergent iterations, indicating good exploration of the parameter space.

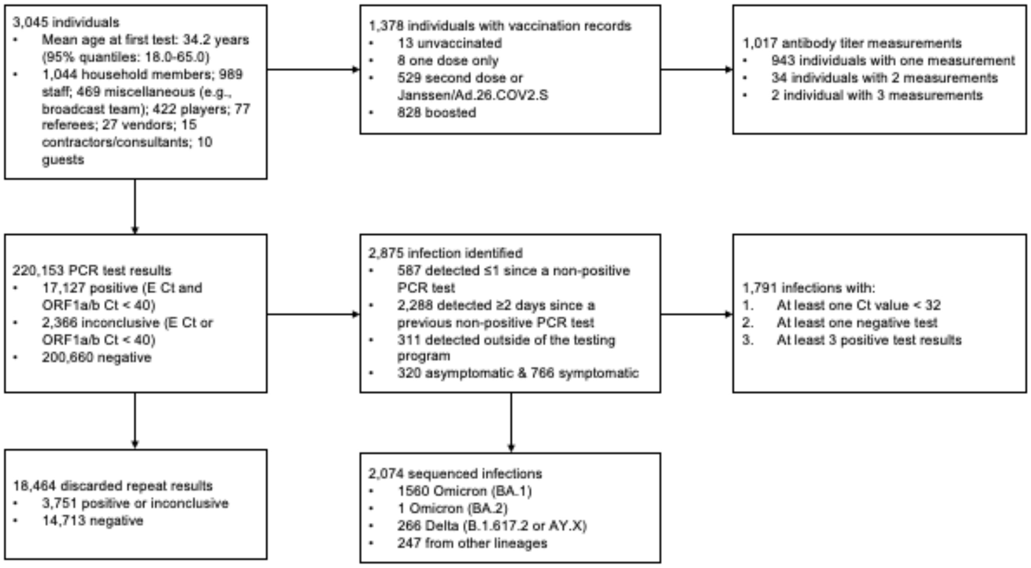


**Figure S1.** Summary of cohort. Top row describes cohort demographics and data on immune histories. Middle row describes infection data. Bottom row provides additional information on the infection data.

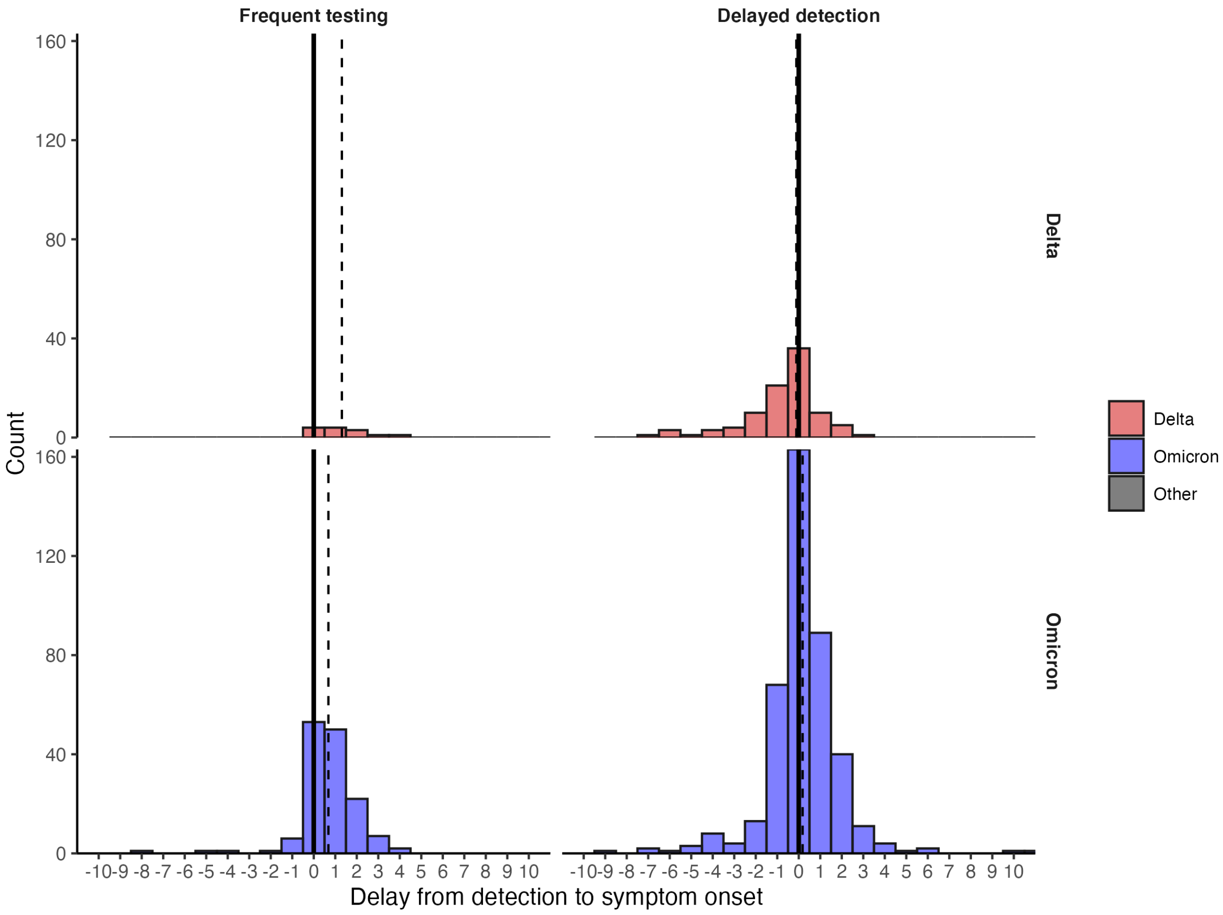


**Figure S2.** Distribution of delays from detection to symptom onset among individuals with known symptom status. Dashed lines mark the median delay between detection and symptom onset. Solid lines mark the day of detection (0).

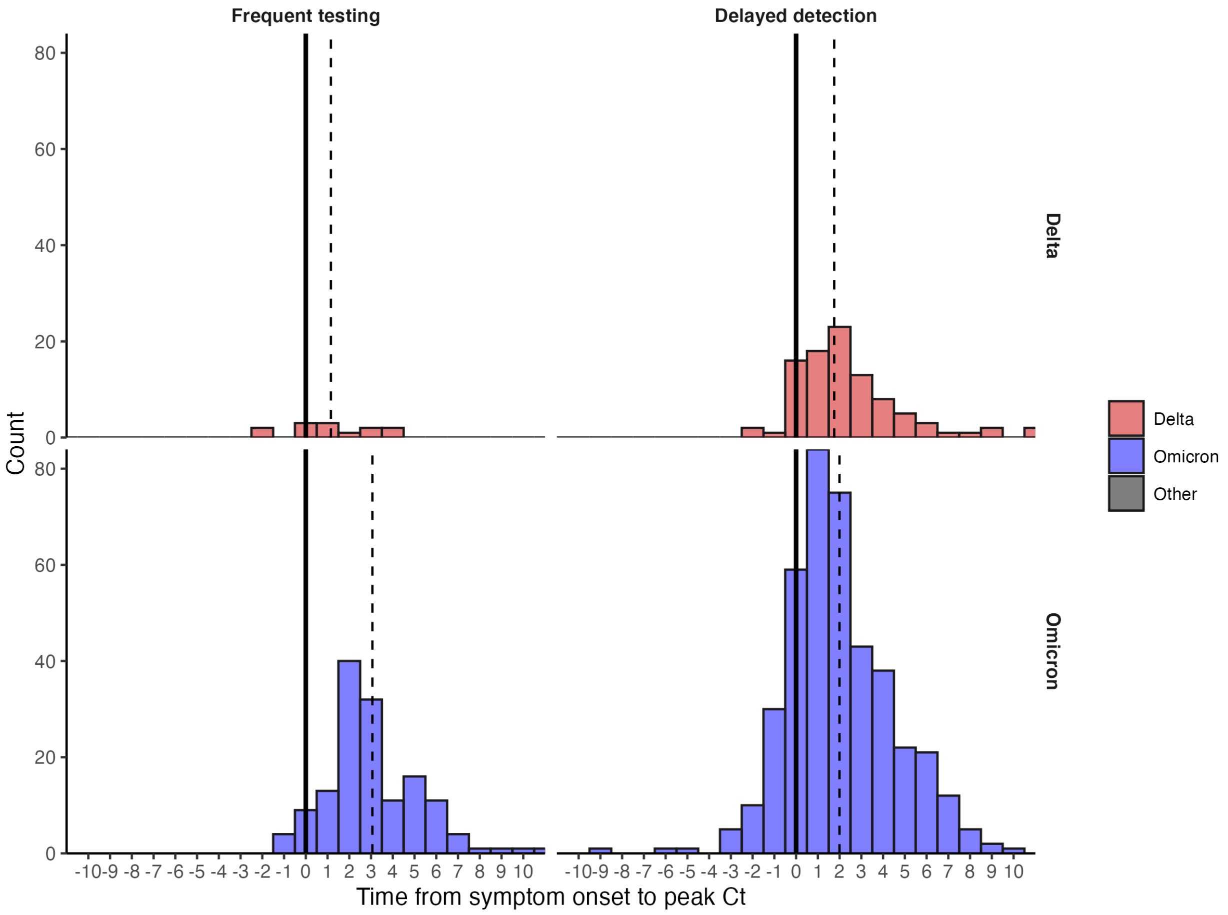


**Figure S3.** Distribution of delays from symptom onset to peak Ct values among individuals with known symptom status. Dashed lines mark the median delay between detection and symptom onset. Solid lines mark the day of symptom onset (0).

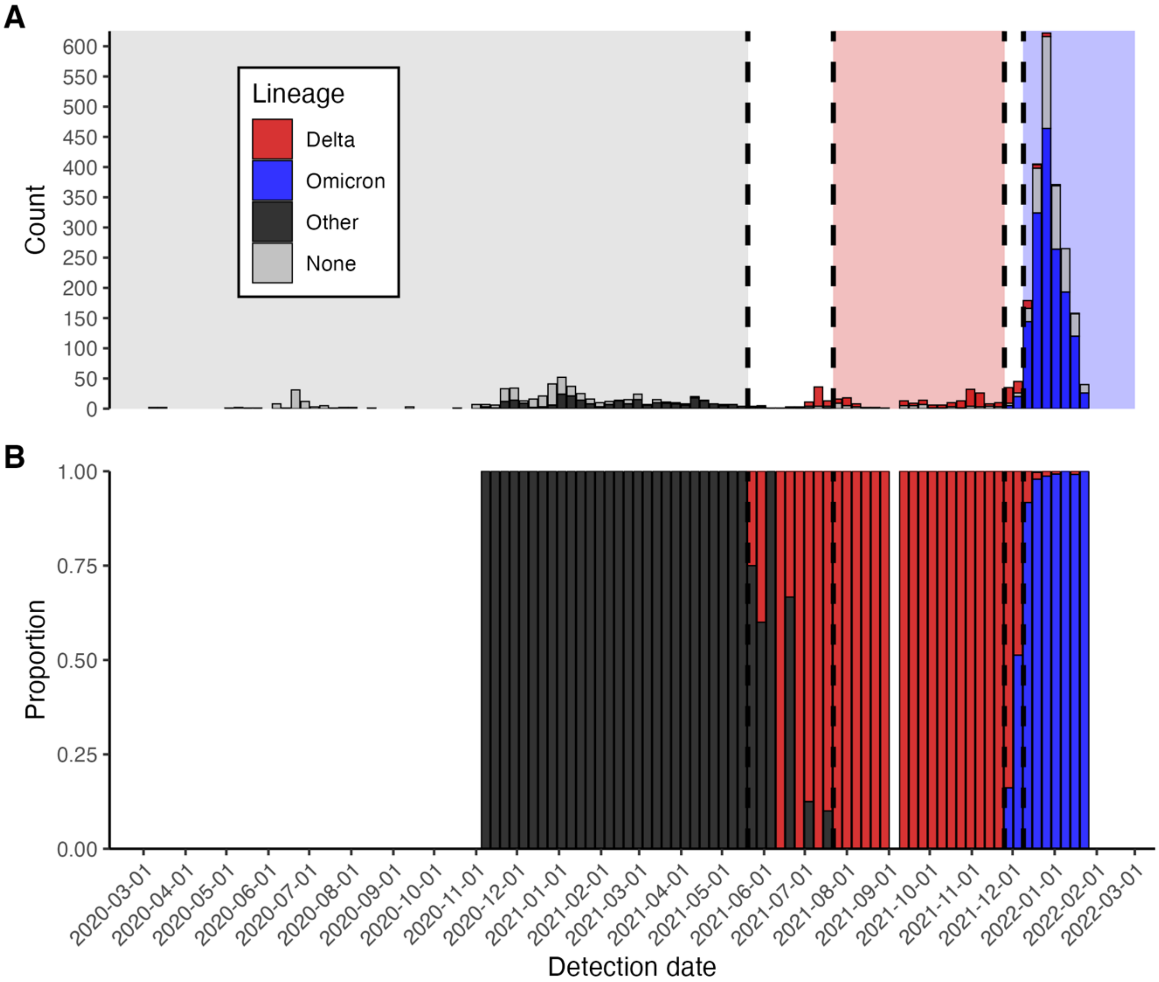


**Figure S4.** (A) Frequency of sequenced and unsequenced detected infections over time by week. Vertical dashed lines and shaded backgrounds demarcate periods of variant dominance. (B) Proportion of sequenced infections attributable to Delta, Omicron or other lineages.

**Table S1.** Number of identified rebounds stratified by variant, either confirmed through sequencing or assumed based on detection date. Rebounds are defined here as any trajectory with an initial Ct value <30, followed by a sequence of two or more consecutive negative tests or tests with Ct value ≥30, and subsequently followed by two or more consecutive tests with Ct value <30.

| Lineage | Rebounds | Total infections | Percentage rebounded |
|---|---|---|---|
| **Omicron** | 36 | 877 | 4.10% |
| **Delta** | 1 | 178 | 0.562% |
| **Other** | 3 | 279 | 1.08% |

**Table S2.** Number of identified rebounds stratified by vaccination status. Rebounds are defined here as any trajectory with an initial Ct value <30, followed by a sequence of two or more consecutive negative tests or tests with Ct value ≥30, and subsequently followed by two or more consecutive tests with Ct value <30.

| Vaccination status | Rebounds | Total infections | Percentage rebounded |
|---|---|---|---|
| **Boosted** | 32 | 494 | 6.48% |
| **No record** | 5 | 398 | 1.26% |
| **Second dose** | 3 | 323 | 0.929% |
| **Unvaccinated** | 2 | 159 | 1.26% |

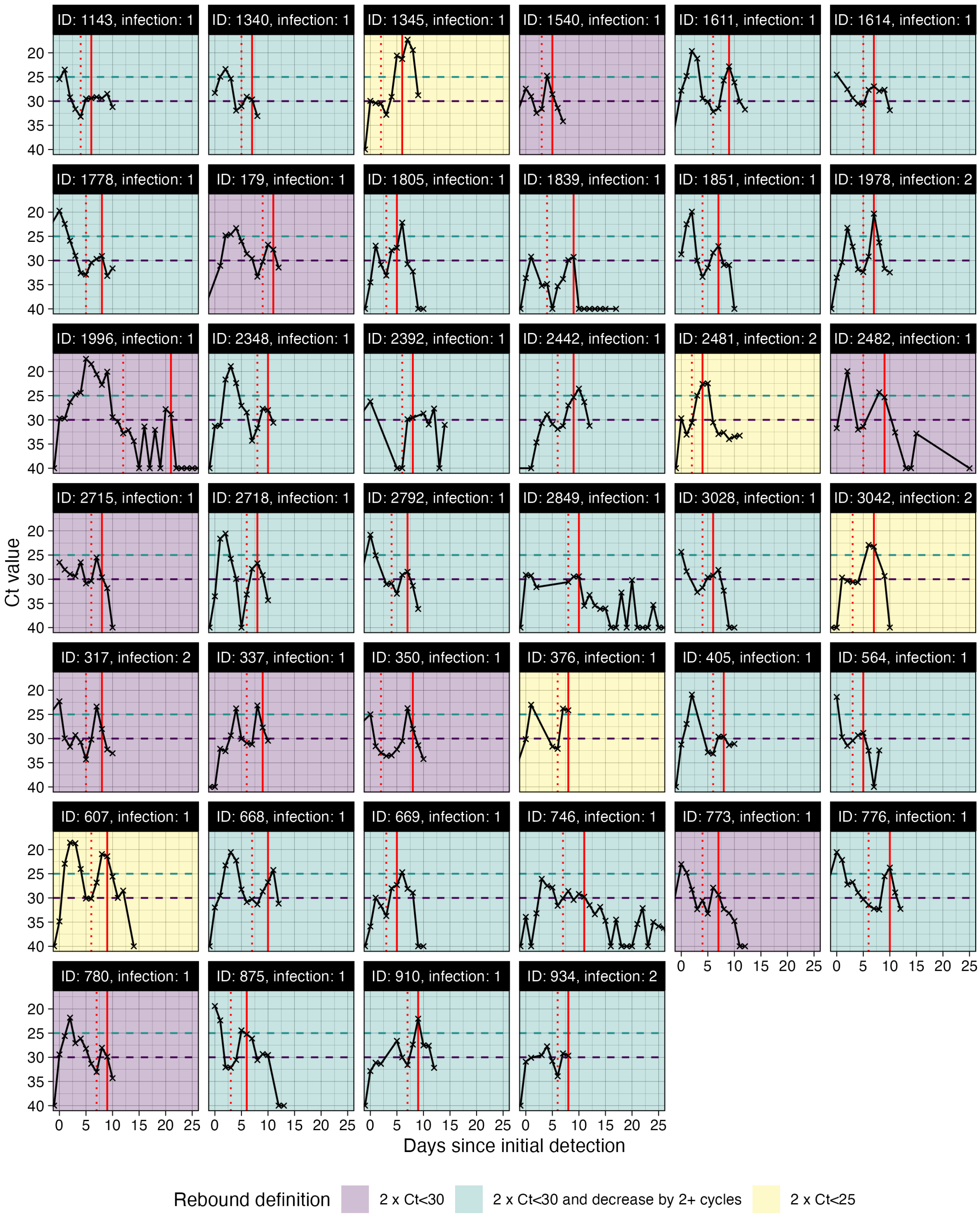


**Supplementary Figure S5. All viral trajectories classified as rebound shown in Figure 1B.** Subplots are colored by the most stringent definition for rebound. To be included here, individuals must have 2+ consecutive days of Ct≥30 after an initial Ct<30. The vertical red dotted line marks this initial clearance time. Trajectories are then classified as rebounds following either two consecutive tests with Ct<30 (purple), two consecutive tests with Ct<30 but with at least a 2 Ct decrease (green), or two consecutive tests with Ct<25 (yellow). The vertical red line marks the timing of rebound detection. The horizontal dashed lines show the different Ct value thresholds for rebound classification. Panels are labeled by arbitrary person ID and infection number.

**Table S3.** Comparison of linear logistic regression models predicting probability of Ct<30 on each day since detection among individuals in the frequent testing group. Models are ranked based on their expected log pointwise predictive density (ELPD), where a lower ELPD implies better prediction accuracy. Model weight refers to the weight of each model in a Bayesian Model Averaging analysis, where a higher value implies a greater contribution to model prediction when combining multiple models. AUC = area under the curve.

| Model | ELPD difference | SE difference | Model weight | AUC | Classification accuracy | Accuracy (>=30) | Accuracy (<30) |
|---|---|---|---|---|---|---|---|
| Cumulative number of exposures and lineage | 0.000 | 0.000 | 0.189 | 0.900 | 0.828 | 0.894 | 0.637 |
| Days since previous exposure and lineage | -6.053 | 11.613 | 0.281 | 0.899 | 0.826 | 0.892 | 0.634 |
| Vaccination status and lineage | -6.907 | 11.836 | 0.292 | 0.899 | 0.826 | 0.887 | 0.649 |
| Cumulative number of exposures | -24.622 | 8.645 | 0.237 | 0.895 | 0.824 | 0.898 | 0.609 |
| Vaccination status | -43.428 | 13.353 | 0.000 | 0.892 | 0.821 | 0.888 | 0.626 |
| Days since previous exposure | -44.910 | 13.731 | 0.001 | 0.893 | 0.821 | 0.890 | 0.619 |
| Lineage | -74.343 | 12.956 | 0.000 | 0.889 | 0.819 | 0.892 | 0.607 |
| Baseline | -80.956 | 14.080 | 0.000 | 0.887 | 0.819 | 0.895 | 0.597 |

**Table S4.** Comparison of linear logistic regression models predicting probability of Ct<30 on each day since detection among individuals in the delayed testing group. Models are ranked based on their expected log pointwise predictive density (ELPD), where a lower ELPD implies better prediction accuracy. Model weight refers to the weight of each model in a Bayesian Model Averaging analysis, where a higher value implies a greater contribution to model prediction when combining multiple models. AUC = area under the curve.

| Model | ELPD difference | SE difference | Model weight | AUC | Classification accuracy | Accuracy (>=30) | Accuracy (<30) |
|---|---|---|---|---|---|---|---|
| Cumulative number of exposures and lineage | 0.000 | 0.000 | 0.314 | 0.912 | 0.848 | 0.895 | 0.696 |
| Cumulative number of exposures | -3.543 | 6.862 | 0.282 | 0.911 | 0.847 | 0.895 | 0.695 |
| Vaccination status and lineage | -19.169 | 12.714 | 0.404 | 0.911 | 0.846 | 0.890 | 0.705 |
| Vaccination status | -31.913 | 13.496 | 0.000 | 0.909 | 0.846 | 0.890 | 0.704 |
| Days since previous exposure and lineage | -44.479 | 12.735 | 0.000 | 0.910 | 0.846 | 0.891 | 0.705 |
| Lineage | -61.323 | 13.566 | 0.000 | 0.908 | 0.844 | 0.883 | 0.719 |
| Days since previous exposure | -87.874 | 15.524 | 0.000 | 0.906 | 0.844 | 0.882 | 0.721 |
| Baseline | -117.764 | 17.502 | 0.000 | 0.904 | 0.840 | 0.877 | 0.724 |

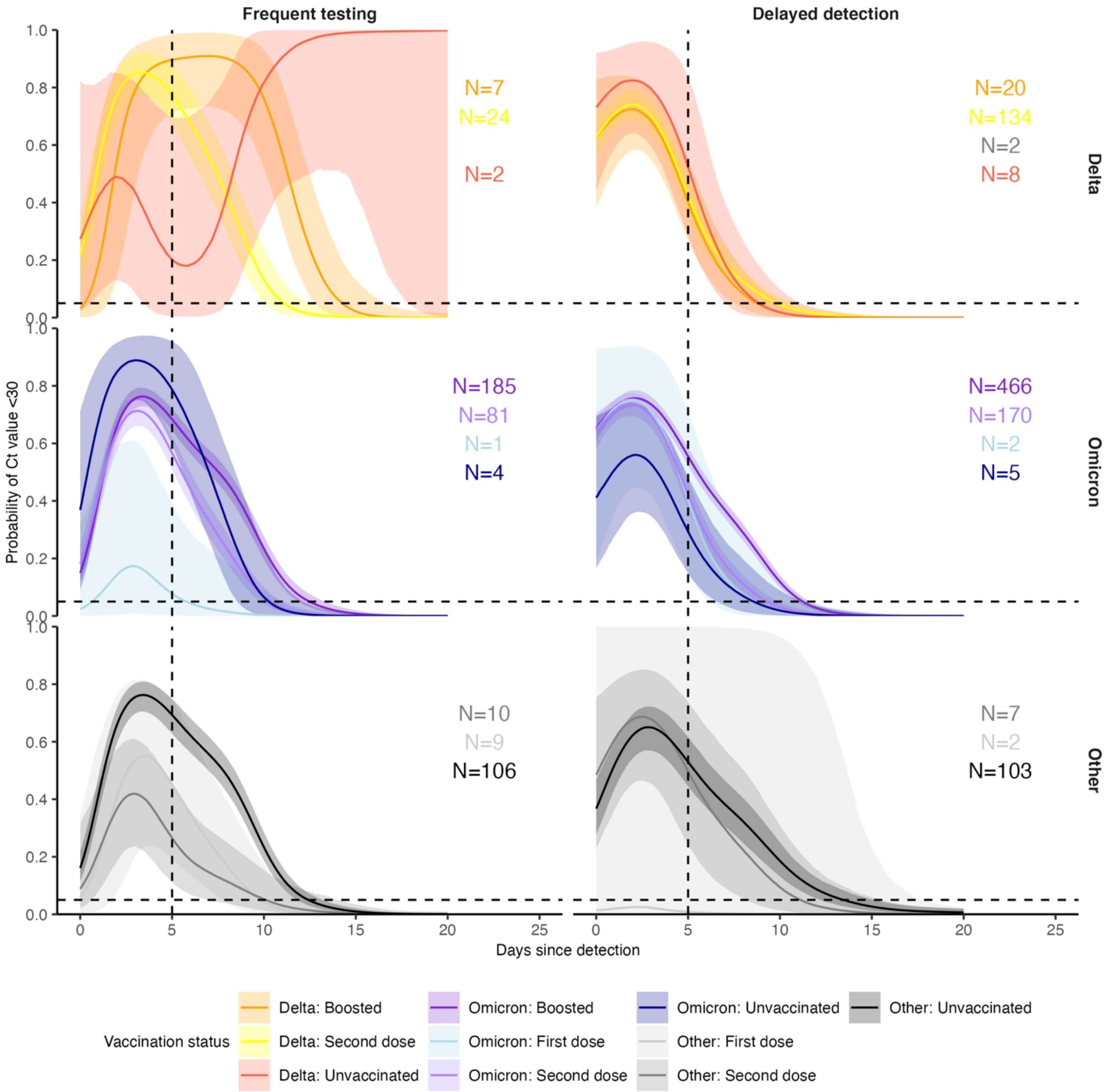


**Figure S6.** Proportion of infections with Ct value <30 on each day post detection by confirmed or suspected variant, vaccination status and detection group. Solid colored lines and shaded ribbons are posterior estimates from a generalized linear model predicting probability of Ct value <30 as a function of days since detection and vaccination status, showing the posterior mean (solid line) and 95% credible intervals (shaded ribbon) of each conditional effect. Dotted horizontal and vertical lines show 5% probability and day 5 post detection respectively.

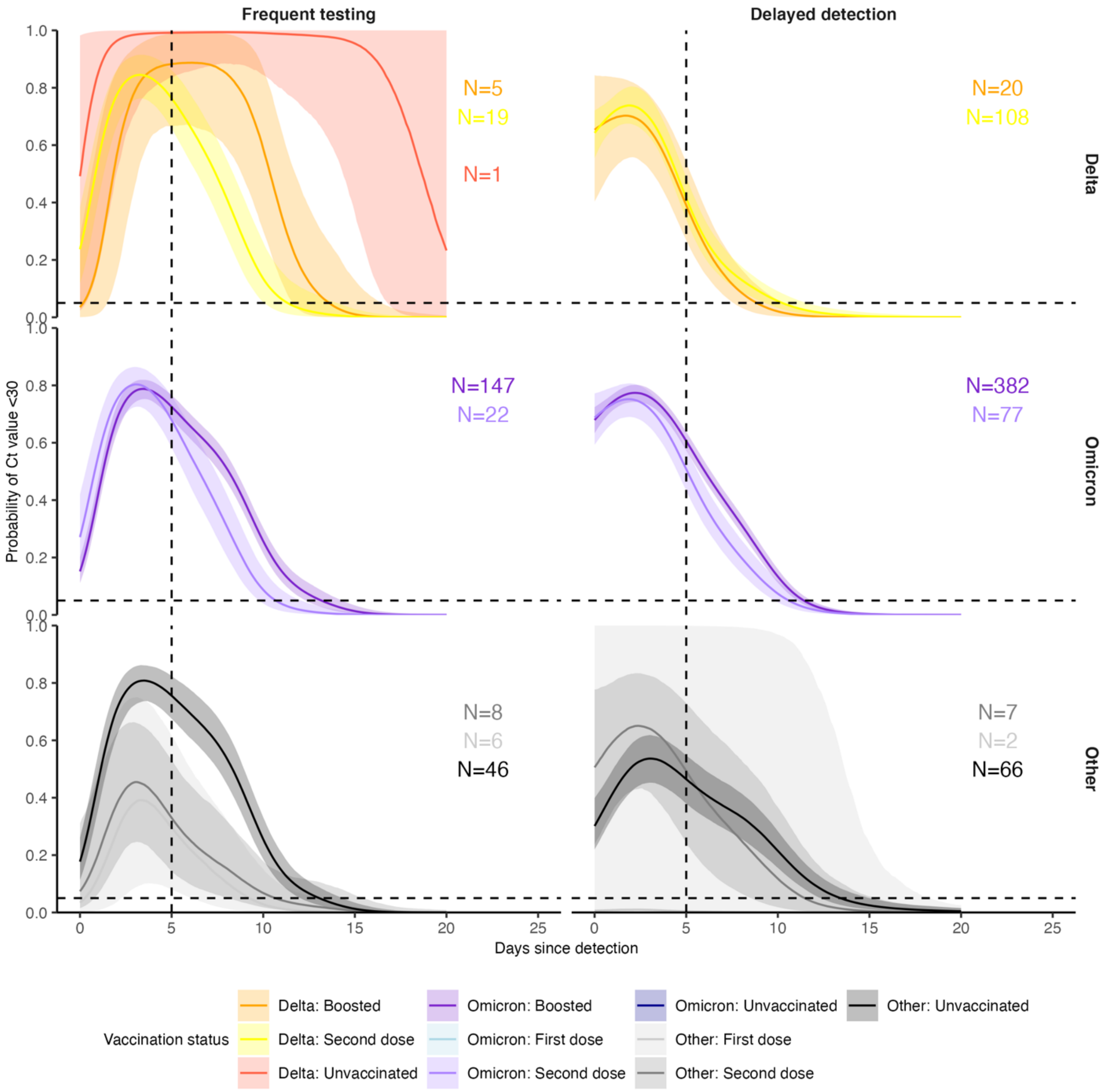


**Figure S7.** Identical to Supplementary Figure 6, but after excluding data from all players.

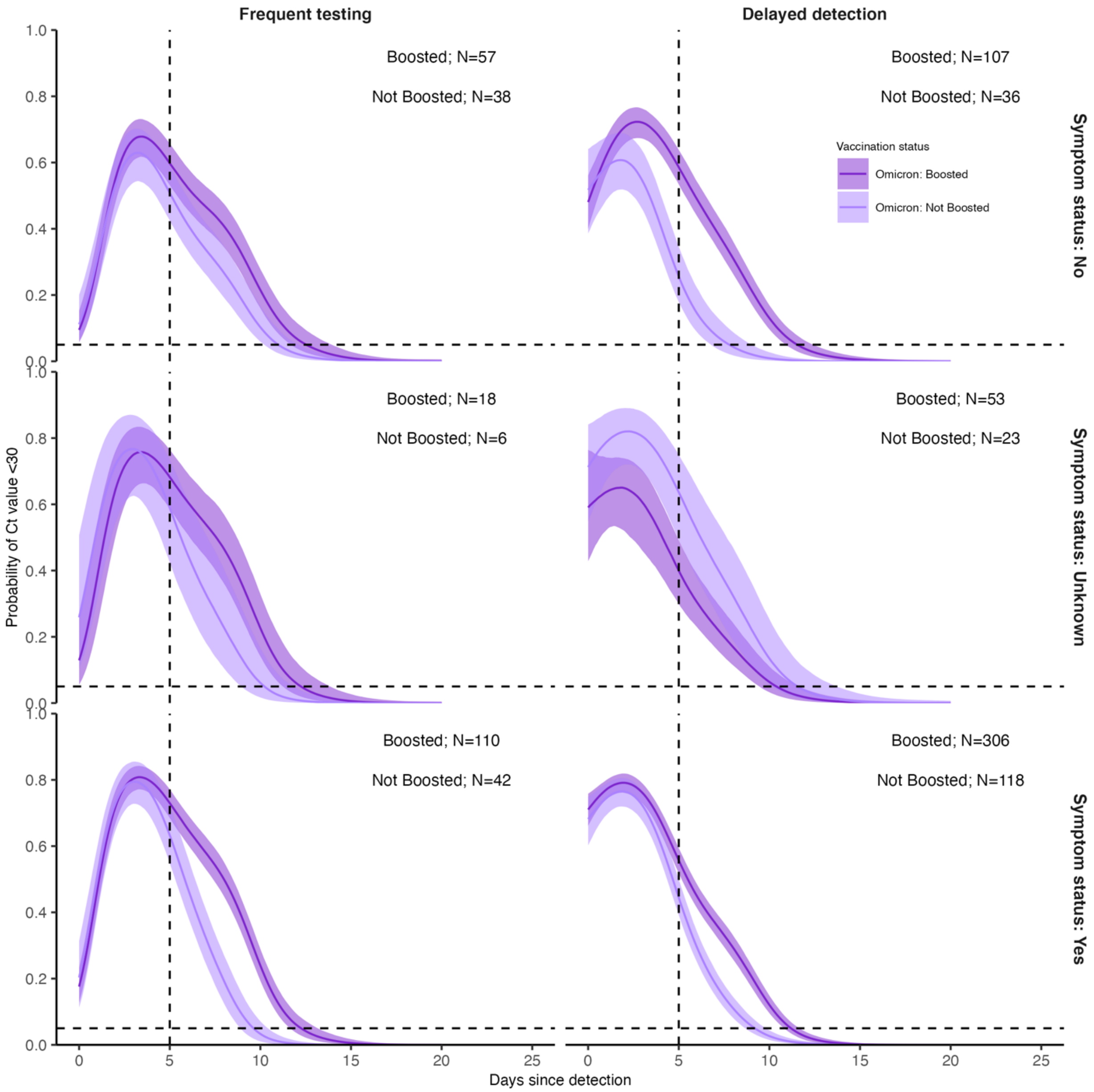


**Figure S8.** Proportion of Omicron infections, stratified by symptom status, with Ct value <30 on each day post detection by booster status and detection group. Solid colored lines and shaded ribbons are posterior estimates from a generalized linear model predicting probability of Ct value <30 as a function of days since detection and vaccination status, showing the posterior mean (solid line) and 95% credible intervals (shaded ribbon) of each conditional effect.

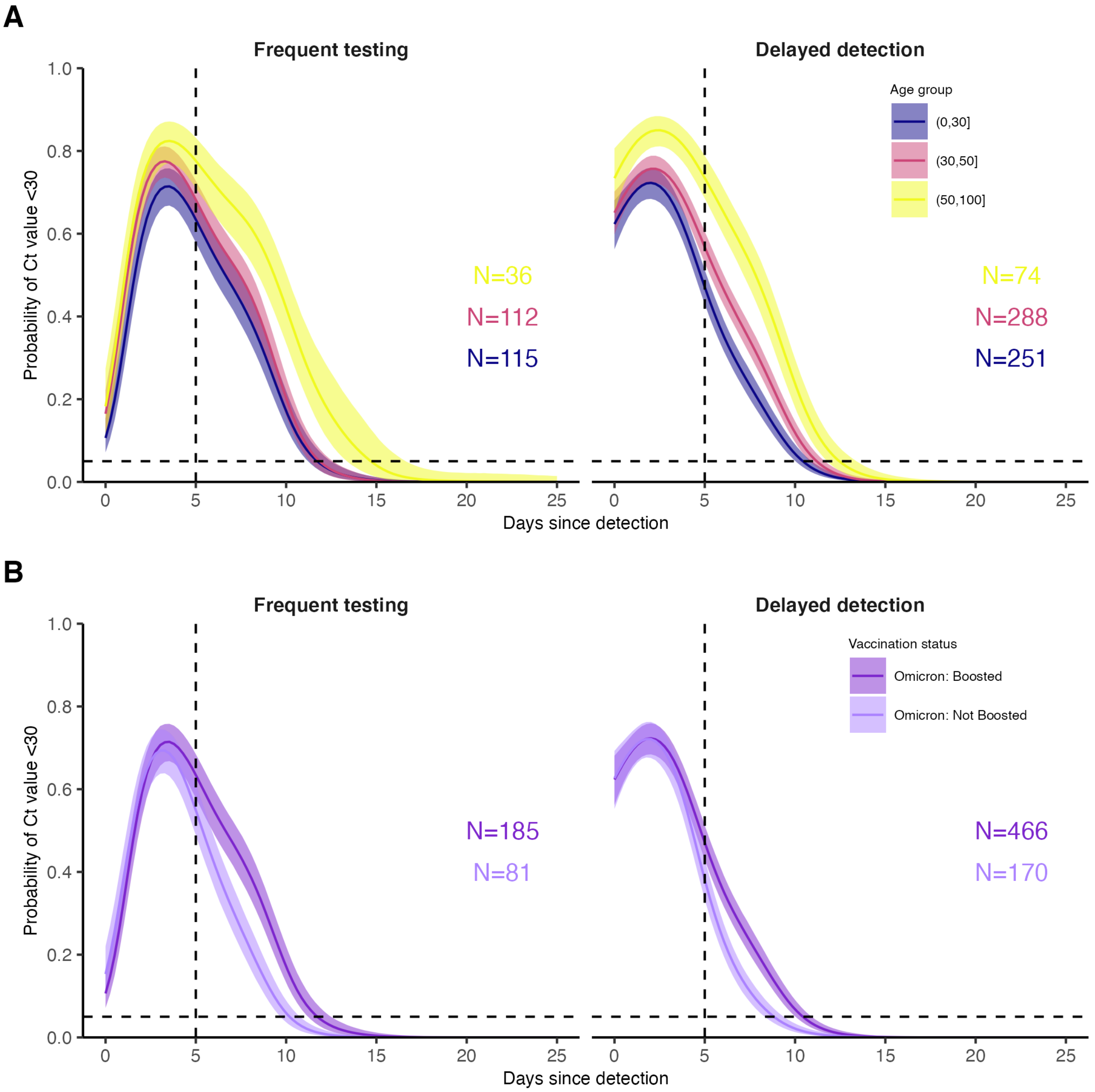


**Figure S9.** Proportion of BA.1-infected individuals with Ct value <30 on each day post detection stratified by detection group and either (**A**) age group after conditioning on vaccination status and lineage (conditioned on BA.1 infection and boosted status) or (**B**) vaccination status after conditioning on age group (conditioned on BA.1 infection and age group <30 years). Posterior estimates are from a generalized linear model predicting probability of Ct value <30 with spline terms for the interaction between days since detection with age group and the interaction between days since detection with vaccination status and variant. Solid colored lines and shaded ribbons show the posterior mean (solid line) and 95% credible intervals (shaded ribbon) of each conditional effect.

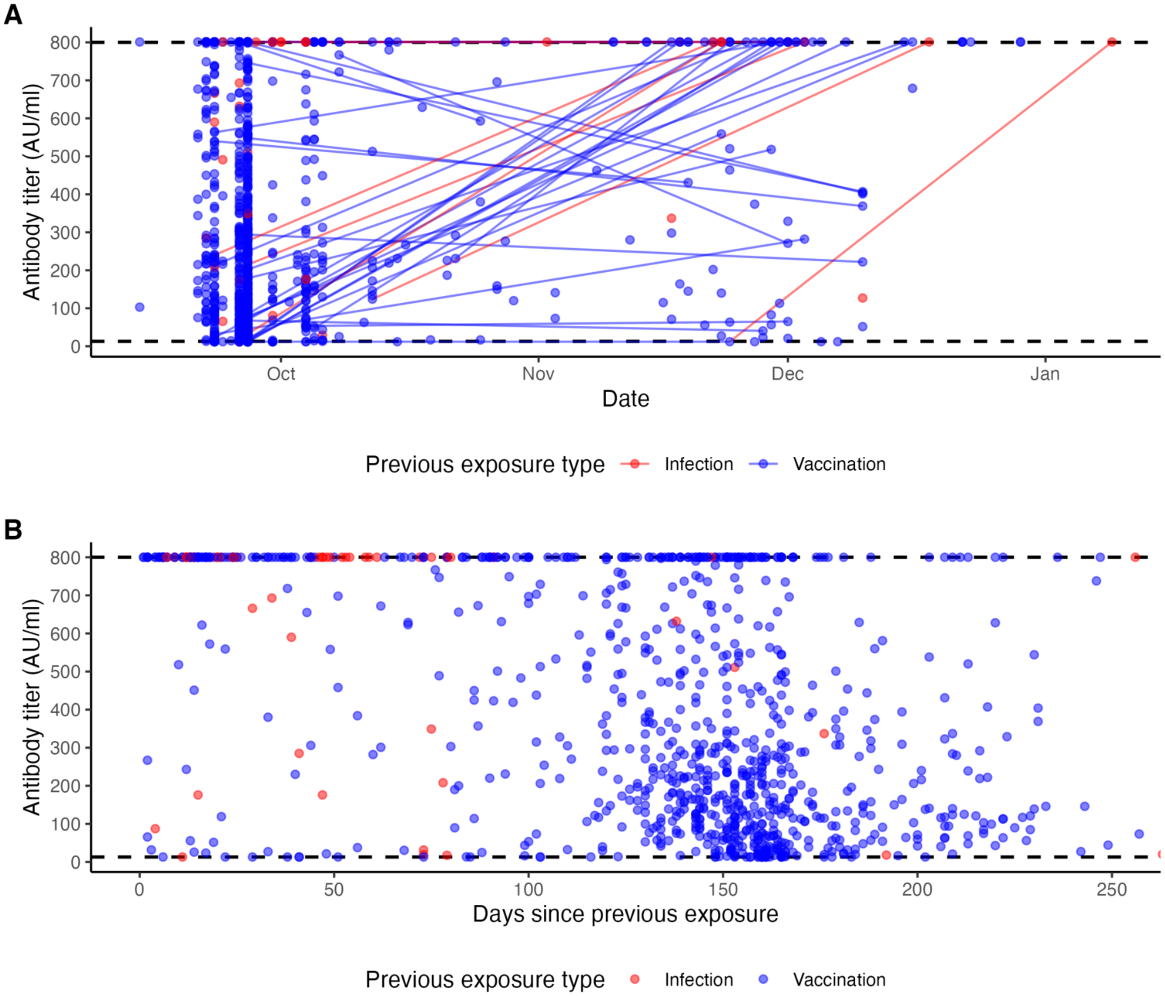


**Figure S10.** (**A**) Measured antibody titers by date of sample collection. Lines show longitudinal samples from the same individual, colored by the most recent exposure at the time of sample collection. Lines going up therefore represent antibody boosting events, and lines going down represent waning. (**B**) Measured antibody titers by days since previous exposure at time of sample collection.

204

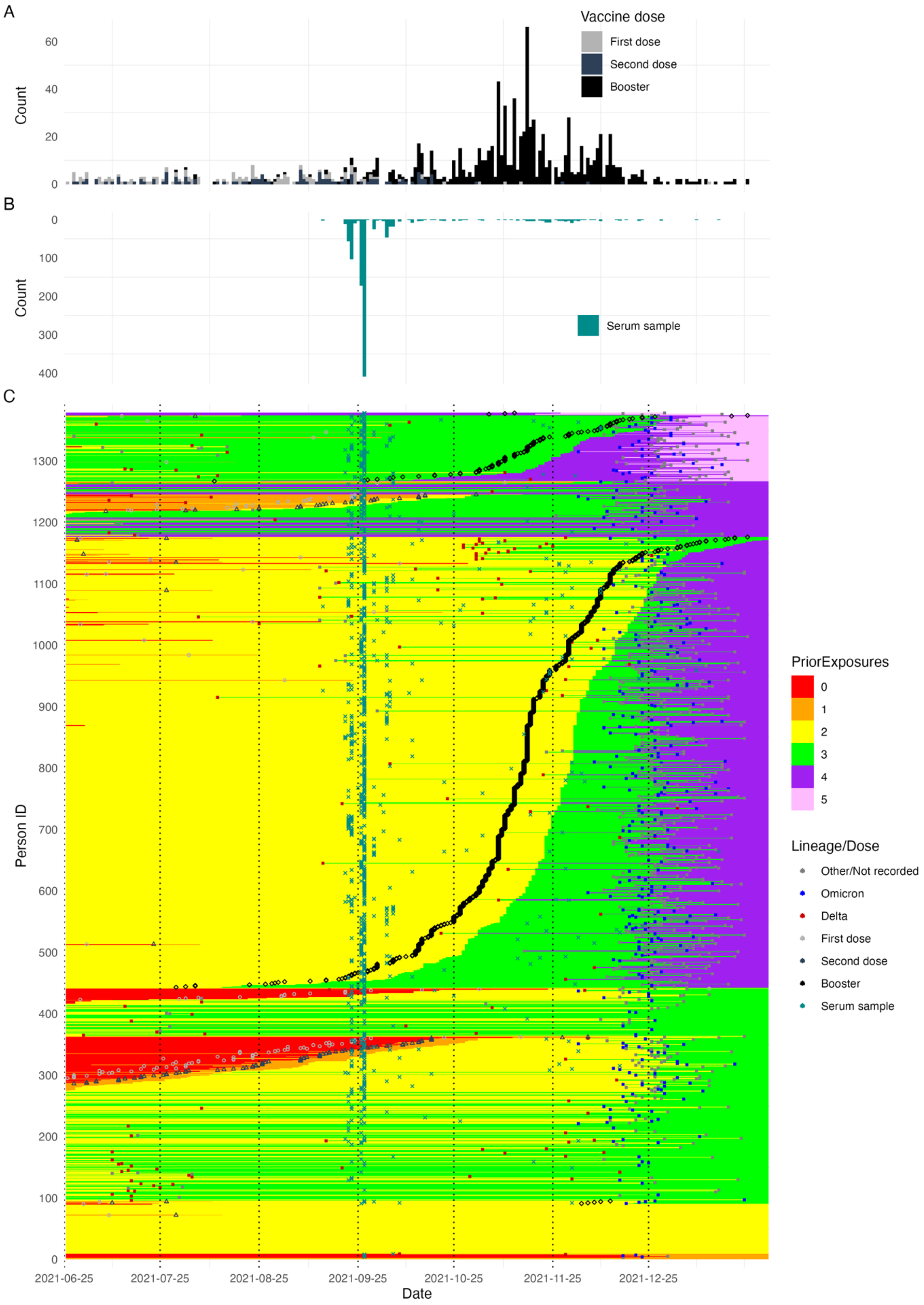

A
Vaccine dose
First dose
Second dose
Booster
Count
60
40
20
0
B
Count
0
100
200
300
400
Serum sample
C
Person ID
1300
1200
1100
1000
900
800
700
600
500
400
300
200
100
0
2021-06-25
2021-07-25
2021-08-25
2021-09-25
2021-10-25
2021-11-25
2021-12-25
Date
PriorExposures
0
1
2
3
4
5
Lineage/Dose
Other/Not recorded
Omicron
Delta
First dose
Second dose
Booster
Serum sample

**Figure S11.** (**A**) Distribution of vaccination dates (note that most first doses were administered prior to 2021-06-25). (**B**) Distribution of serum sample times. (**C**) Heatmap of individual exposure status over time. Rows represent individuals and columns represent date. Each cell is shaded by the number of prior exposures at that date. Points show detected infections, recorded vaccinations, and serum samples.

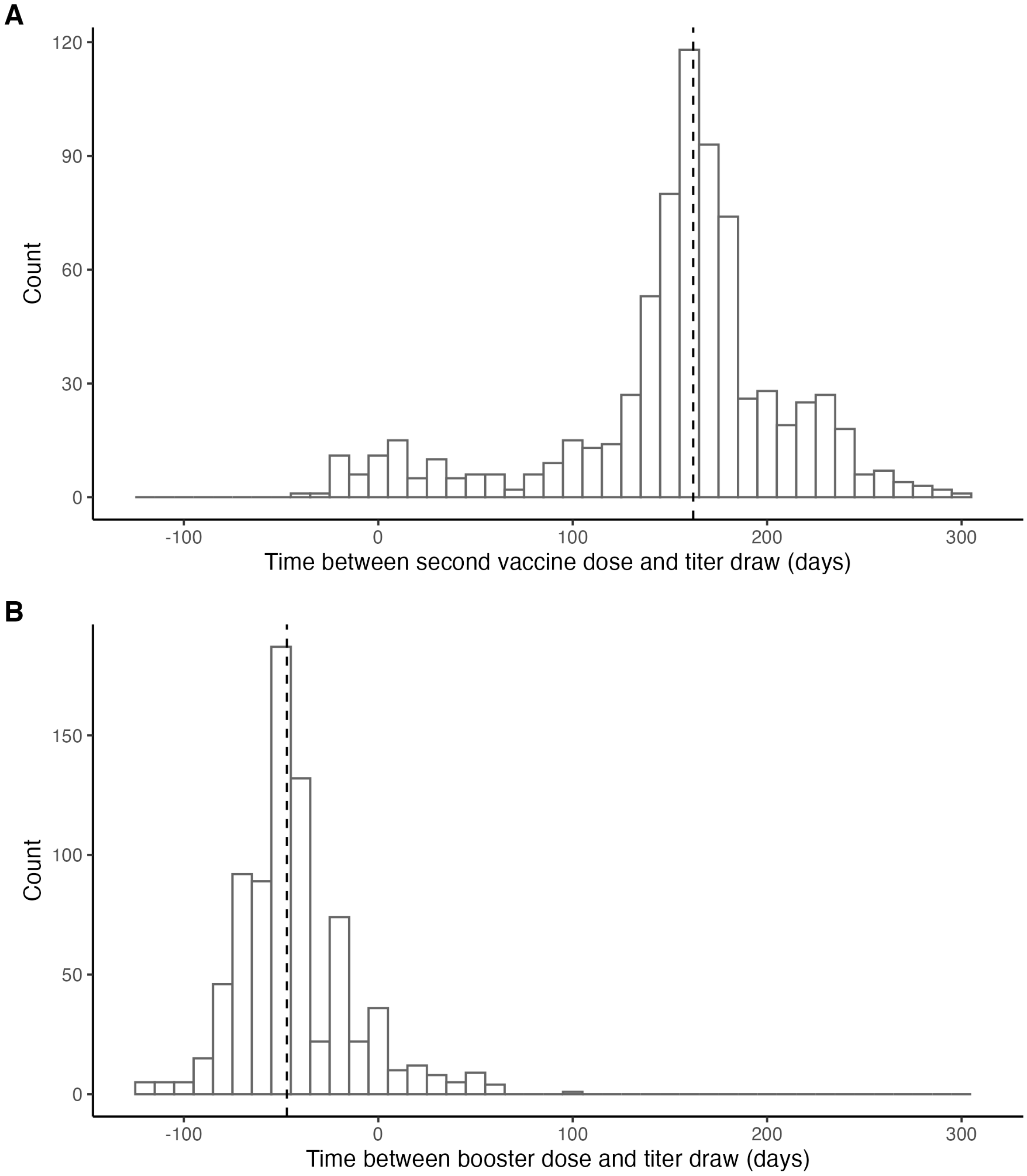


**Figure S12. Histogram of time between (A) second vaccine dose and antibody titer measurement and (B) booster dose and antibody titer measurement.** Dashed line marks the median lag (162 days). 1 individual was infected between receiving their second vaccine dose and having a titer measurement taken (Delta infection). 42 individuals were infected between having their titer measurement taken and receiving their booster vaccine dose (32 Delta; 9 unsequenced; 1 confirmed Omicron BA.1).

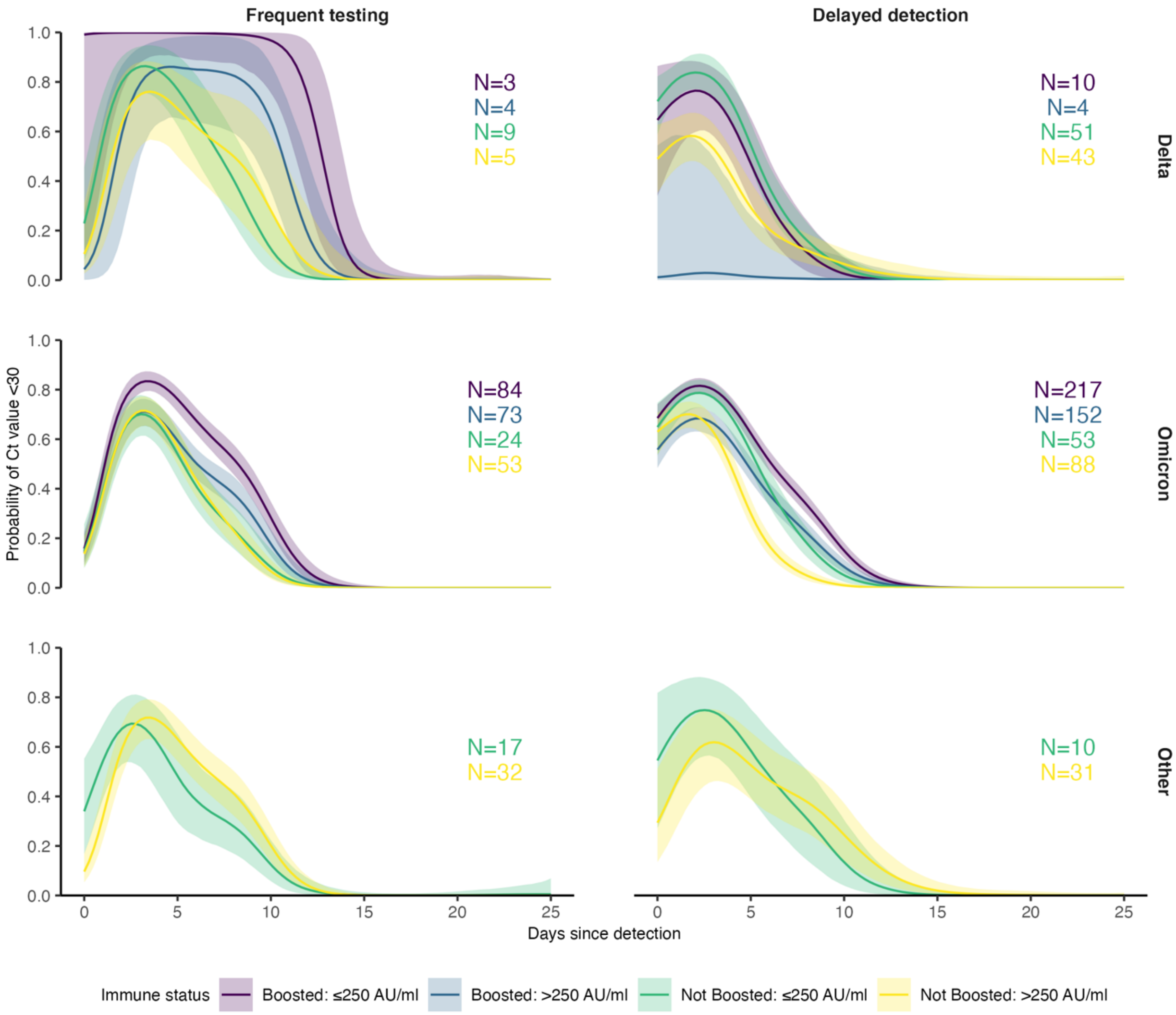


**Figure S13.** Proportion of infections with Ct value <30 on each day post detection stratified by single point-in-time anti-spike antibody titer against ancestral SARS-CoV-2 measured by the Diasorin Trimeric Assay. Solid colored lines and shaded ribbons are posterior estimates from a generalized linear model predicting probability of Ct value <30 as a function of days since detection and titer/vaccination status category, showing the posterior mean (solid line) and 95% credible intervals (shaded ribbon) of each conditional effect.

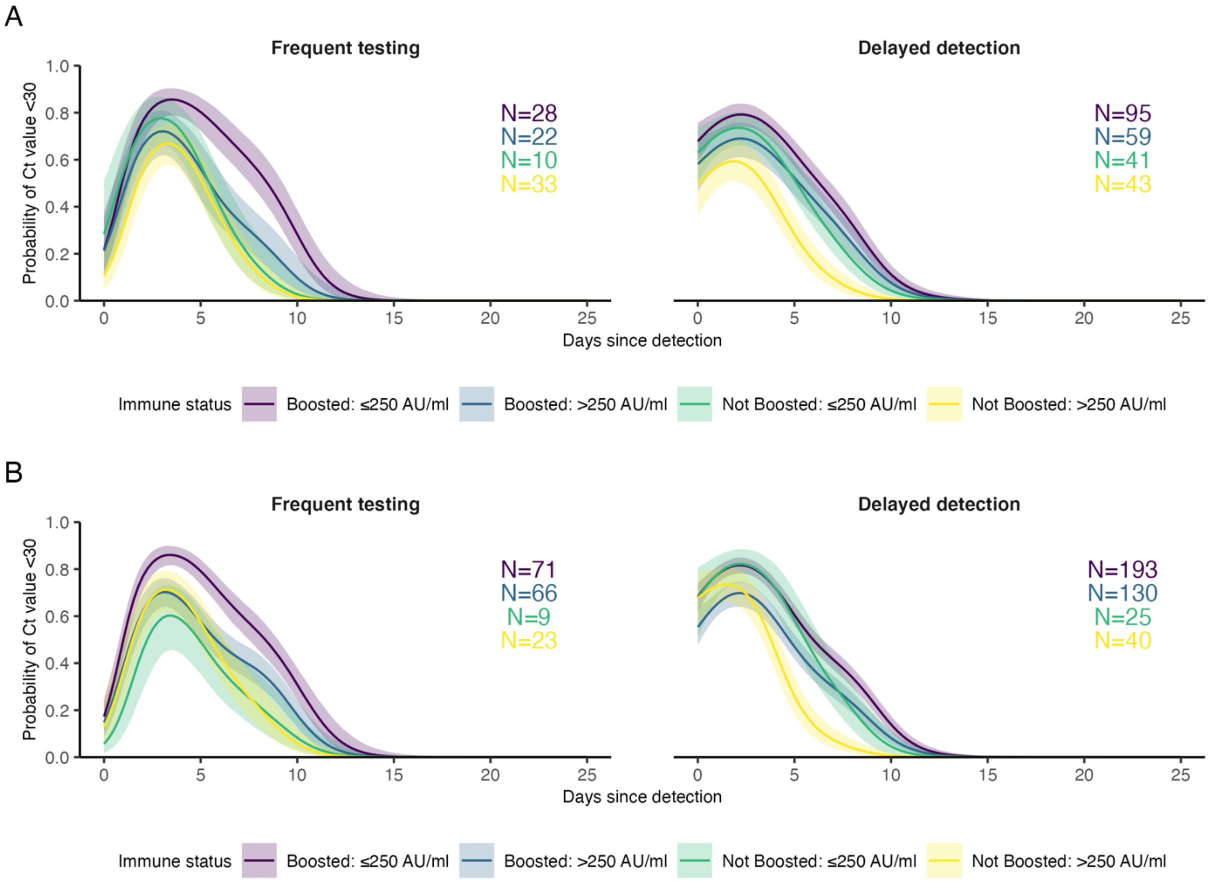


**Figure S14.** Identical to Omicron plots shown in Supplementary Figure 11, but only including individuals who had antibody titers measured between 100 and 200 days following a known previous infection of vaccination (A) or including infections between 60 and 90 days after an antibody titer measurement (B).

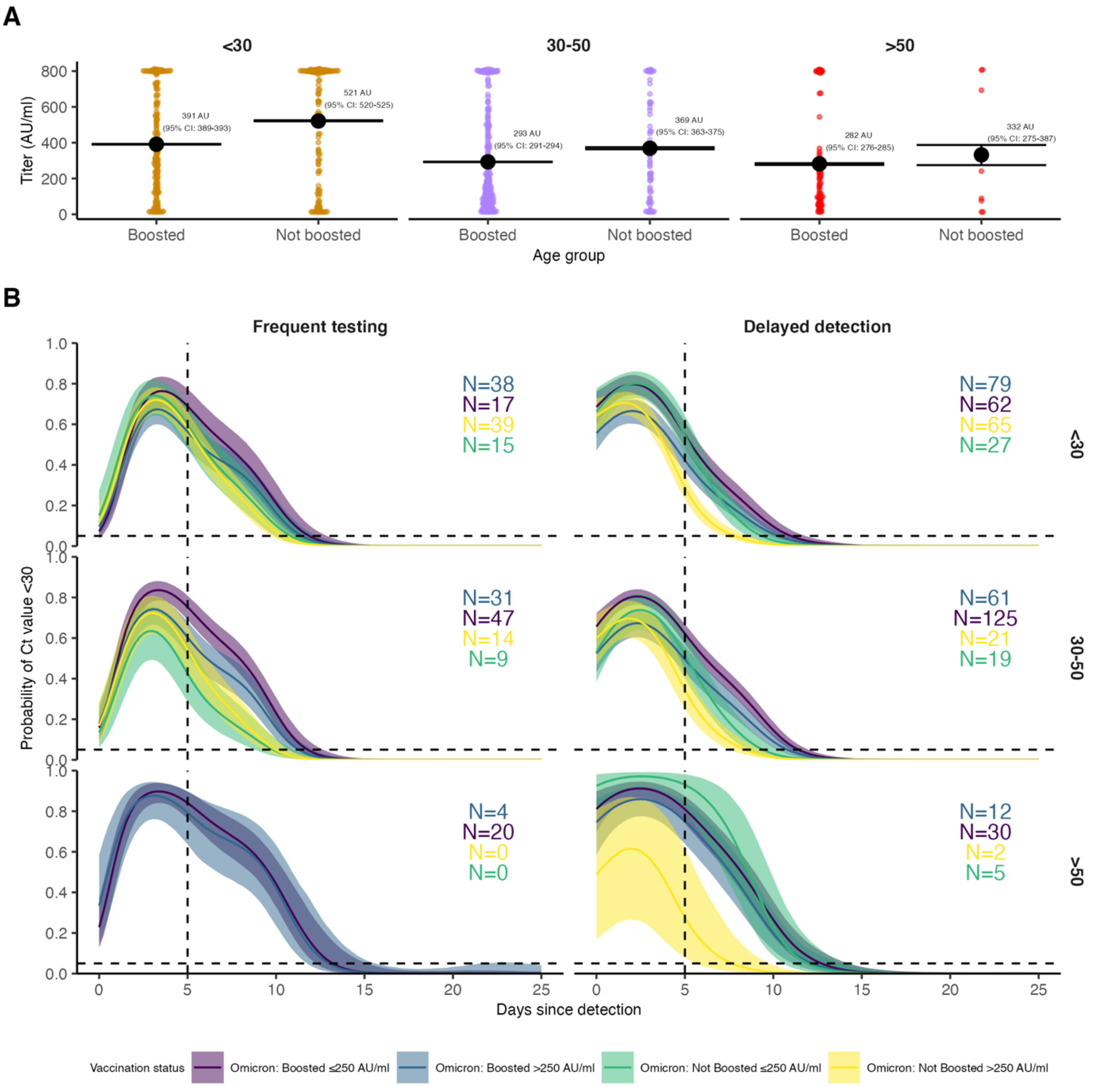


**Figure S15.** (**A**) Distribution of antibody titers among Omicron BA.1-infected individuals (colored points) stratified by age group and vaccination status, with mean titers (large black points) and bootstrapped 95% confidence intervals for the mean (horizontal lines). Note that stratification is by infection and not individual, and that antibody titers were measured at a single point in time rather than near the time of infection. The Diasorin Trimeric Assay values are truncated between 13 and 800 AU/ml. (**B**) Proportion of BA.1-infected individuals with Ct value <30 on each day post detection stratified by detection group after conditioning on age group. Posterior estimates are from a generalized linear model predicting probability of Ct value <30 with spline terms for the interaction between days since detection with age group and the interaction between days since detection with vaccination status and antibody titer group. Solid colored lines and shaded ribbons show the posterior mean (solid line) and 95% credible intervals (shaded ribbon) of each conditional effect.

**Table S5. Posterior estimates of viral trajectory attributes by variant and vaccination status.** Estimates are posterior means with 95% credible intervals.

| Trajectory value | Variant/vaccination status | Estimate |
|---|---|---|
| Peak viral load (Ct) | Other: Unvaccinated | 25.0 (24.2, 25.9) |
| | Delta: 1-2 doses | 22.4 (21.4, 23.5) |
| | Omicron: 1-2 doses | 25.6 (25.0, 26.2) |
| | Omicron: Boosted | 25.7 (25.4, 26.1) |
| Peak viral load ($log_{10}$ copies/ml) | Other: Unvaccinated | 6.8 (6.6, 7.0) |
| | Delta: 1-2 doses | 7.5 (7.2, 7.8) |
| | Omicron: 1-2 doses | 6.6 (6.5, 6.8) |
| | Omicron: Boosted | 6.6 (6.5, 6.7) |
| Proliferation time (days) | Other: Unvaccinated | 3.5 (3.1, 3.9) |
| | Delta: 1-2 doses | 3.6 (3.1, 4.1) |
| | Omicron: 1-2 doses | 3.6 (3.3, 4.0) |
| | Omicron: Boosted | 4.0 (3.8, 4.3) |
| Clearance time (days) | Other: Unvaccinated | 9.9 (9.2, 10.6) |
| | Delta: 1-2 doses | 7.6 (7.0, 8.3) |
| | Omicron: 1-2 doses | 6.2 (5.8, 6.6) |
| | Omicron: Boosted | 8.4 (8.0, 8.7) |

**Table S6. Posterior estimates of viral trajectory attributes by variant and antibody titer.** Estimates are posterior means with 95% credible intervals.

| Trajectory attribute | Variant/immune status | Estimate |
|---|---|---|
| Peak viral load (Ct) | Other: Unexposed | 24.8 (24.0, 25.6) |
| | Delta: Exposed, ≤250 AU | 22.1 (20.6, 23.4) |
| | Delta: Exposed, >250 AU | 24.4 (22.7, 26.1) |
| | Omicron: Exposed, ≤250 AU | 25.2 (24.7, 25.6) |
| | Omicron: Exposed, >250 AU | 26.2 (25.7, 26.6) |
| Peak viral load ($\log_{10}$ copies/ml) | Other: Unexposed | 6.9 (6.6, 7.1) |
| | Delta: Exposed, ≤250 AU | 7.6 (7.3, 8.0) |
| | Delta: Exposed, >250 AU | 7.0 (6.5, 7.4) |
| | Omicron: Exposed, ≤250 AU | 6.8 (6.7, 6.9) |
| | Omicron: Exposed, >250 AU | 6.5 (6.4, 6.6) |
| Proliferation time (days) | Other: Unexposed | 3.5 (3.2, 3.9) |
| | Delta: Exposed, ≤250 AU | 3.6 (2.8, 4.5) |
| | Delta: Exposed, >250 AU | 4.2 (3.3, 5.3) |
| | Omicron: Exposed, ≤250 AU | 3.9 (3.6, 4.2) |
| | Omicron: Exposed, >250 AU | 3.7 (3.5, 4.0) |
| Clearance time (days) | Other: Unexposed | 9.8 (9.1, 10.6) |
| | Delta: Exposed, ≤250 AU | 7.7 (6.9, 8.7) |
| | Delta: Exposed, >250 AU | 7.6 (6.4, 8.8) |
| | Omicron: Exposed, ≤250 AU | 8.4 (8.0, 8.8) |
| | Omicron: Exposed, >250 AU | 6.9 (6.6, 7.3) |

**Table S7. Posterior estimates of Omicron BA.1 viral trajectory attributes by symptom and vaccination status.** Estimates are posterior means with 95% credible intervals.

| Trajectory attribute | Variant/immune status | Estimate |
|---|---|---|
| Peak viral load (Ct) | Omicron: 1-2 doses, no symptoms | 26.7 (25.7, 27.7) |
| | Omicron: 1-2 doses, symptoms | 25.3 (24.5, 26.1) |
| | Omicron: boosted, no symptoms | 26.3 (25.7, 27) |
| | Omicron: boosted, symptoms | 25.4 (25, 25.8) |
| Peak viral load ($log_{10}$ copies/ml) | Omicron: 1-2 doses, no symptoms | 6.3 (6.1, 6.6) |
| | Omicron: 1-2 doses, symptoms | 6.7 (6.5, 6.9) |
| | Omicron: boosted, no symptoms | 6.4 (6.3, 6.6) |
| | Omicron: boosted, symptoms | 6.7 (6.6, 6.8) |
| Proliferation time (days) | Omicron: 1-2 doses, no symptoms | 3.7 (3.2, 4.2) |
| | Omicron: 1-2 doses, symptoms | 3.7 (3.2, 4.2) |
| | Omicron: boosted, no symptoms | 4.3 (3.9, 4.8) |
| | Omicron: boosted, symptoms | 3.8 (3.5, 4.1) |
| Clearance time (days) | Omicron: 1-2 doses, no symptoms | 5.9 (5.2, 6.6) |
| | Omicron: 1-2 doses, symptoms | 6.3 (5.8, 6.8) |
| | Omicron: boosted, no symptoms | 7.4 (6.8, 8.0) |
| | Omicron: boosted, symptoms | 8.7 (8.3, 9.1) |

**Table S8. Posterior estimates of Omicron BA.1 viral trajectory attributes by symptom and antibody titer.** Estimates are posterior means with 95% credible intervals.

| Trajectory attribute | Variant/immune status | Estimate |
|---|---|---|
| Peak viral load (Ct) | Omicron: Low titer, no symptoms | 26.1 (25.2, 26.9) |
| | Omicron: Low titer, symptoms | 24.8 (24.3, 25.4) |
| | Omicron: High titer, no symptoms | 26.7 (25.9, 27.5) |
| | Omicron: High titer, symptoms | 25.9 (25.4, 26.5) |
| Peak viral load ($log_{10}$ copies/ml) | Omicron: Low titer, no symptoms | 6.5 (6.3, 6.8) |
| | Omicron: Low titer, symptoms | 6.9 (6.7, 7.0) |
| | Omicron: High titer, no symptoms | 6.3 (6.1, 6.6) |
| | Omicron: High titer, symptoms | 6.6 (6.4, 6.7) |
| Proliferation time (days) | Omicron: Low titer, no symptoms | 4.3 (3.8, 4.9) |
| | Omicron: Low titer, symptoms | 3.6 (3.3, 4.0) |
| | Omicron: High titer, no symptoms | 3.8 (3.4, 4.3) |
| | Omicron: High titer, symptoms | 3.8 (3.4, 4.2) |
| Clearance time (days) | Omicron: Low titer, no symptoms | 7.4 (6.7, 8.1) |
| | Omicron: Low titer, symptoms | 8.7 (8.2, 9.3) |
| | Omicron: High titer, no symptoms | 6.4 (5.8, 7.1) |
| | Omicron: High titer, symptoms | 7.1 (6.6, 7.5) |

**Table S9. Posterior estimates of Omicron BA.1 viral RNA clearance times by age and vaccination status.** Estimates are posterior means with 95% credible intervals. Low titer is ≤250 AU, high titer is >250 AU.

| **Trajectory attribute** | **Variant/immune status** | **Estimate** |
|---|---|---|
| Peak viral load (Ct) | Omicron: <30, unboosted | 25.6 (25, 26.3) |
| | Omicron: <30, boosted | 25.8 (25.1, 26.4) |
| | Omicron: 30-50, unboosted | 25.3 (24.2, 26.5) |
| | Omicron: 30-50, boosted | 25.7 (25.2, 26.2) |
| | Omicron: 50+, unboosted | 25.5 (22.8, 27.9) |
| | Omicron: 50+, boosted | 25.5 (24.6, 26.3) |
| Peak viral load ($\log_{10}$ copies/ml) | Omicron: <30, unboosted | 6.6 (6.4, 6.8) |
| | Omicron: <30, boosted | 6.6 (6.4, 6.8) |
| | Omicron: 30-50, unboosted | 6.7 (6.4, 7) |
| | Omicron: 30-50, boosted | 6.6 (6.5, 6.7) |
| | Omicron: 50+, unboosted | 6.7 (6, 7.4) |
| | Omicron: 50+, boosted | 6.7 (6.5, 6.9) |
| Proliferation time (days) | Omicron: <30, unboosted | 3.7 (3.4, 4.1) |
| | Omicron: <30, boosted | 4.2 (3.8, 4.7) |
| | Omicron: 30-50, unboosted | 3.6 (3, 4.4) |
| | Omicron: 30-50, boosted | 4 (3.6, 4.3) |
| | Omicron: 50+, unboosted | 4.4 (3, 6.3) |
| | Omicron: 50+, boosted | 3.9 (3.4, 4.4) |
| Clearance time (days) | Omicron: <30, unboosted | 6 (5.6, 6.5) |
| | Omicron: <30, boosted | 7.2 (6.7, 7.7) |
| | Omicron: 30-50, unboosted | 6.4 (5.7, 7.2) |
| | Omicron: 30-50, boosted | 8.6 (8.1, 9.1) |
| | Omicron: 50+, unboosted | 8.7 (6.6, 11.2) |
| | Omicron: 50+, boosted | 9.6 (8.8, 10.6) |

**Table S10. Posterior estimates of Omicron BA.1 viral RNA clearance times by age and titer.** Estimates are posterior means with 95% credible intervals. Low titer is ≤250 AU, high titer is >250 AU.

| Trajectory attribute | Variant/immune status | Estimate |
|---|---|---|
| Peak viral load (Ct) | Omicron: <30, low titer | 24.8 (24, 25.5) |
| | Omicron: <30, high titer | 26.2 (25.6, 26.8) |
| | Omicron: 30-50, low titer | 25.4 (24.8, 26) |
| | Omicron: 30-50, high titer | 26.1 (25.4, 26.8) |
| | Omicron: 50+, low titer | 24.9 (23.8, 25.9) |
| | Omicron: 50+, high titer | 26.3 (24.8, 27.6) |
| Peak viral load ($\log_{10}$ copies/ml) | Omicron: <30, low titer | 6.9 (6.7, 7.1) |
| | Omicron: <30, high titer | 6.5 (6.3, 6.7) |
| | Omicron: 30-50, low titer | 6.7 (6.5, 6.9) |
| | Omicron: 30-50, high titer | 6.5 (6.3, 6.7) |
| | Omicron: 50+, low titer | 6.8 (6.6, 7.1) |
| | Omicron: 50+, high titer | 6.5 (6.1, 6.9) |
| Proliferation time (days) | Omicron: <30, low titer | 3.9 (3.4, 4.3) |
| | Omicron: <30, high titer | 4 (3.6, 4.4) |
| | Omicron: 30-50, low titer | 4 (3.6, 4.4) |
| | Omicron: 30-50, high titer | 3.6 (3.2, 4) |
| | Omicron: 50+, low titer | 3.8 (3.2, 4.5) |
| | Omicron: 50+, high titer | 3.7 (2.9, 4.7) |
| Clearance time (days) | Omicron: <30, low titer | 7.2 (6.7, 7.8) |
| | Omicron: <30, high titer | 6.2 (5.8, 6.7) |
| | Omicron: 30-50, low titer | 8.7 (8.1, 9.3) |
| | Omicron: 30-50, high titer | 7.4 (6.8, 8) |
| | Omicron: 50+, low titer | 9.9 (8.7, 11.1) |
| | Omicron: 50+, high titer | 9.5 (8, 11.1) |

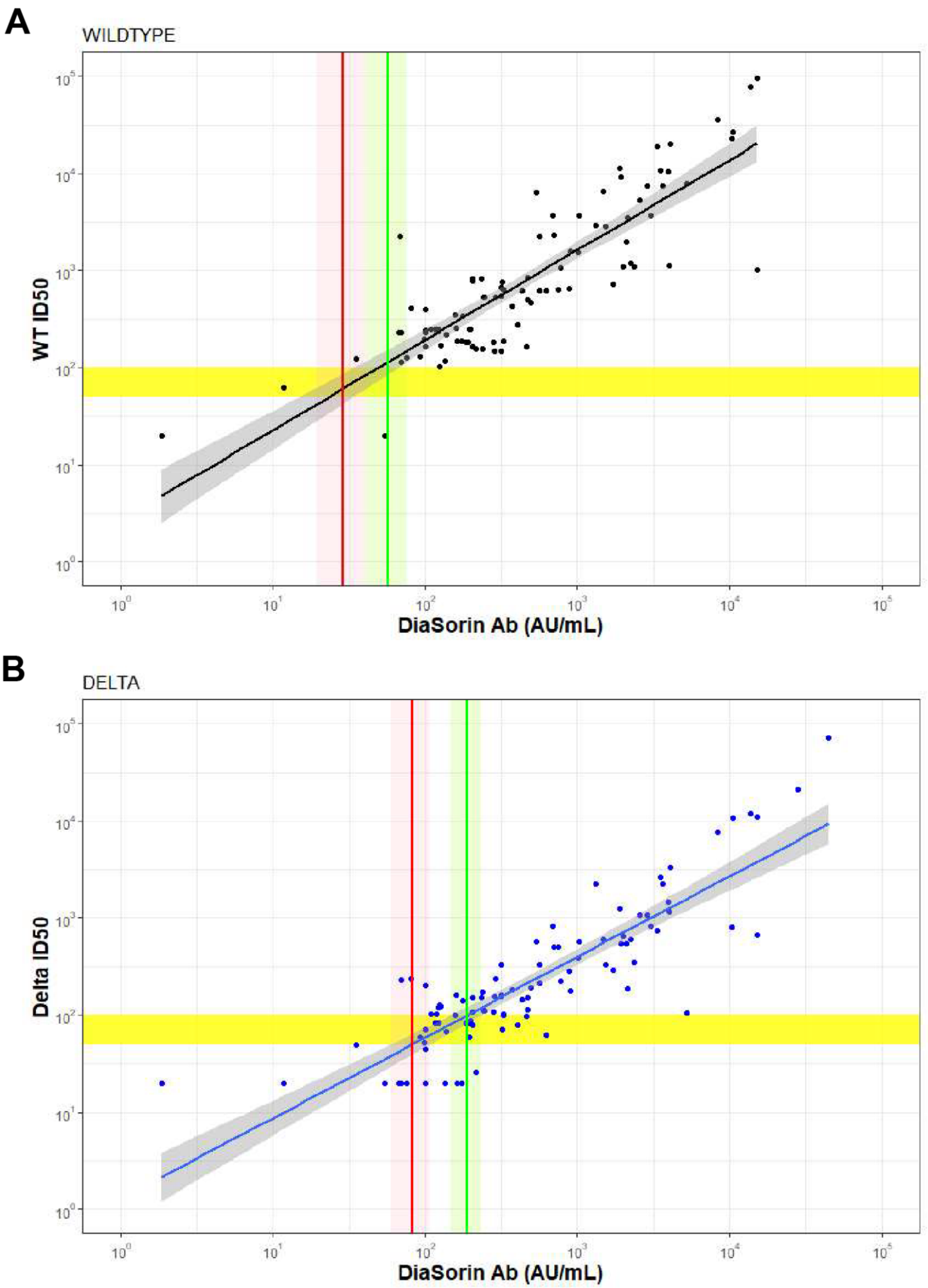


**Figure S16.** Correlation between authentic virus neutralization assay (ID50) and the Diasorin antibody titer against (A) wildtype and (B) Delta. Horizontal yellow bar shows an ID50 titer of 50 and 100 respectively, Diagonal lines and shaded regions show mean and 95% confidence intervals (CI) for a linear regression between the Diason antibody titer and ID50 titer. Vertical line and shaded regions show point estimate and 95% CI for the Diasorin antibody titer corresponding to an ID50 titer of 50 (red) and 100 (green).

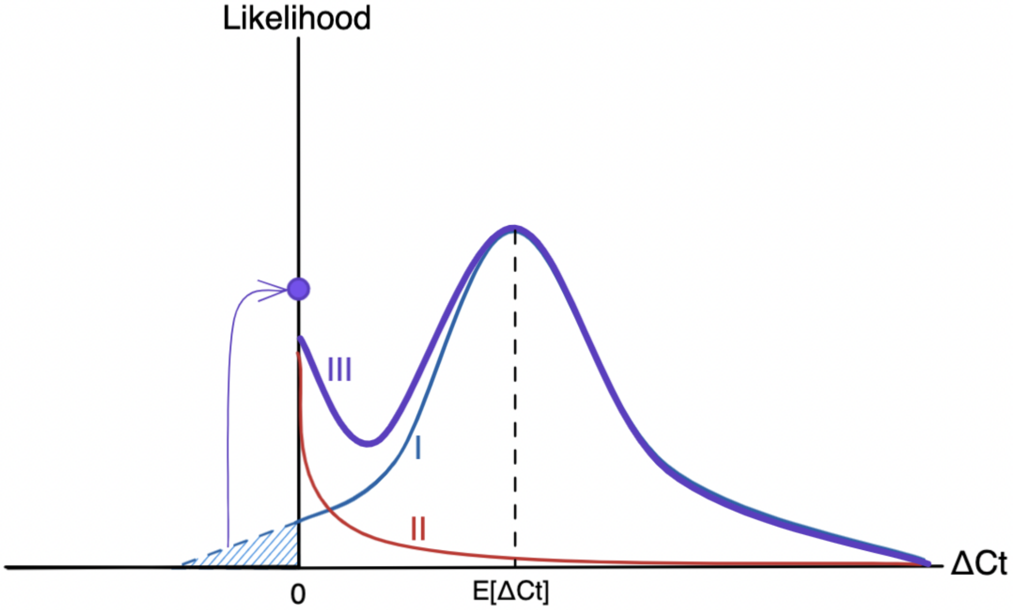


**Figure S17. Schematic diagram of the likelihood function for viral kinetic inference.** The plot depicts the likelihood as a function of ΔCt, the difference between the observed Ct value and the limit of detection, so that ΔCt = 0 (the origin) represents observations at the limit of detection, with viral load increasing toward the right-hand side of the plot. The likelihood function (III, purple) is made up of two fundamental components: the process likelihood (I, blue) and the false negative distribution (II, red). The main component of the process likelihood (I) is defined by a normal distribution with mean E[ΔCt], a function of the estimated viral kinetic parameters as defined by the viral kinetic model. Any mass of the process likelihood that extends below the limit of detection (blue hatched region) is instead added to a point probability mass at the origin, since viral loads below the limit of detection register at the limit of detection. The false negative distribution (II) is an exponential distribution with fixed rate to account for a small amount of noise near the limit of detection. Summing the process likelihood (I) and the false negative likelihood (I) using the mixing probability λ (= 1 - sensitivity) yields the overall likelihood (III).
